# Acidity of expiratory aerosols controls the infectivity of airborne influenza virus and SARS-CoV-2

**DOI:** 10.1101/2022.03.14.22272134

**Authors:** Beiping Luo, Aline Schaub, Irina Glas, Liviana K. Klein, Shannon C. David, Nir Bluvshtein, Kalliopi Violaki, Ghislain Motos, Marie Pohl, Walter Hugentobler, Athanasios Nenes, Ulrich K. Krieger, Silke Stertz, Thomas Peter, Tamar Kohn

## Abstract

Enveloped viruses are prone to inactivation when exposed to strong acidity levels characteristic of atmospheric aerosol. Yet, the acidity of expiratory aerosol particles and its effect on airborne virus persistence has not been examined. Here, we combine pH-dependent inactivation rates of influenza A virus and SARS-CoV-2 with microphysical properties of respiratory fluids using a biophysical aerosol model. We find that particles exhaled into indoor air become mildly acidic (pH ≈ 4), rapidly inactivating influenza A virus within minutes, whereas SARS-CoV-2 requires days. If indoor air is enriched with non-hazardous levels of nitric acid, aerosol pH drops by up to 2 units, decreasing 99%-inactivation times for both viruses in small aerosol particles to below 30 seconds. Conversely, unintentional removal of volatile acids from indoor air by filtration may elevate pH and prolong airborne virus persistence. The overlooked role of aerosol pH has profound implications for virus transmission and mitigation strategies.

## Introduction

Respiratory viral infections pose a great burden on human health. An average of 400,000 deaths are associated with influenza globally each year (1), and the ongoing COVID-19 pandemic has already resulted in several million deaths and countless cases of long COVID around the world. To curb the public health and economic impacts of these diseases, health care policy aims to minimize virus transmission. Increasing evidence points to expiratory aerosol particles (see (2) for clarification of terminology) as vehicles for the transmission of influenza virus and SARS-CoV-2 (3). The persistence of these viruses in aerosols is still subject to scientific debate, but it is undisputed that rapid inactivation would contribute to limiting their spread.

Prior studies have investigated the effect of ambient conditions on the inactivation rates of aerosolized respiratory viruses including influenza virus (4–9), SARS-CoV-2 (10–12), and the common cold human coronavirus HCoV-229E (13). Relative humidity (RH) and temperature were the primary variables modulated in these works, with low (∼ 20%), medium (40-60%), and high (65-90%) RH compared at a few select temperatures. Some of these studies identified a ‘U-shaped’ curve of inactivation as a function of RH (4, 7), and it has been suggested that RH affects virus inactivation by controlling evaporation of water from the aerosol particle, thus governing the concentration of inactivation-catalysing solutes (14–16). Beyond this, the mechanism(s) of virus inactivation in aerosol particles remain largely speculative.

A potentially powerful, yet understudied driver of airborne virus inactivation is the aerosol pH. It is established now that aerosol particles can be highly acidic (17), and that some enveloped viruses, including influenza virus, are sensitive to low pH (18). Nevertheless, though previously hypothesized to be a determinant of virus fate (19), the pH of expiratory aerosol particles, and hence its contribution to the inactivation of airborne viruses, remains unknown. The aerosol pH depends on the composition of the aerosol particle and the surrounding air, and it is well characterized for particulate matter equilibrated with inorganic acids and bases (20). Some studies have investigated the role of matrix composition on virus inactivation in particles, including its protective properties (7, 8, 21). However, the impact of air composition beyond RH has been overlooked by scientists to date. To the best of our knowledge, the only attempt to inactivate airborne viruses by - likely inadvertently - modulating aerosol pH is the use of acetic acid from boiling vinegar during the 2002/03 outbreak of SARS-CoV-1 (see (22) and Supplementary Text).

Outdoor airborne particulate matter is often highly acidic, with pH values ranging between -1 and +5 (17, 20). Contrary to expectations, the strength of the acid or base contained in aerosols (expressed by its dissociation constants) may not be the dominant parameter controlling aerosol pH. Rather, the volatility of species is of importance. For example, strong organic acids like HCOOH and CH_3_COOH partition negligibly to aerosol and bear a minor impact on aerosol pH for most atmospherically relevant conditions (23). In contrast, HNO_3_ and NH_3_ partition into aerosol particles and impact pH, albeit buffered by the formation of ammonium nitrate.

Indoor aerosol particles have a variety of sources, including outdoor air, human transpiration and respiration, and building materials. Indoor air tends to have lower levels of gas-phase inorganic acids (e.g., HNO_3_) than outdoor air, owing to their condensation on aerosol particles as well as their efficient removal via deposition on surfaces. Human activities are a source of organic acids and NH_3_ (20, 24, 25), often elevating their levels compared to outdoors. The ratio of indoor to outdoor concentrations is typically 0.1-0.5 for HNO_3_ and 3-30 for NH_3_, causing the pH of indoor aerosol particles to increase compared to outdoor levels. Operation of humidification, ventilation, and air conditioning (HVAC) systems also affect air composition (26) and, hence, likely the pH of indoor aerosol particles. While many outdoor and indoor aerosol particles are in equilibrium with their environment, this can only be expected for exhaled aerosol if given enough time. In the interim, freshly exhaled aerosol can change its pH considerably.

Exhaled air, before mixing into the indoor air, contains high concentrations of ammonia and is characterized by very high concentrations of CO_2_ and high number densities of expiratory aerosol particles. These particles are emitted by breathing, talking, coughing or sneezing, and contain a complex aqueous mixture of ions, proteins and surfactants. Although the pH of exhaled breath condensate has been investigated (27), there is no study that quantifies the pH of respiratory aerosol - especially when it equilibrates with the acidic or alkaline gases present in the indoor air within a few seconds to minutes of exhalation.

Here, we investigate the role of aerosol acidity in the inactivation of airborne influenza A virus (IAV) and two coronaviruses, SARS-CoV-2 and HCoV-229E in indoor environments. We accomplish this in three steps by first determining the pH-dependent inactivation kinetics of IAV, SARS-CoV-2 and HCoV-229E in bulk samples of representative respiratory fluids, then measuring the thermodynamic and kinetic properties of microscopic particles of these fluids, and finally jointly applying the inactivation kinetics and aerosol properties in a biophysical model to determine inactivation in the aerosol system. We then use the model to investigate the possibility of using gaseous nitric acid (HNO_3_) in indoor environments at non-hazardous concentrations to lower the pH of respiratory aerosol for a wide range of sizes, and thus to effectively reduce the risk of transmission.

## Results and Discussion

### Kinetics of pH-mediated inactivation of influenza virus and coronavirus

Inactivation kinetics of IAV (strain A/WSN/33), SARS-CoV-2 (BetaCoV/Germany/BavPat1/2020) and HCoV-229E (strain HCoV-229E-Ren) were determined over a pH range from neutral to strongly acidic, after immersion in bulk solutions of synthetic lung fluid (SLF; see Table S1 for composition), mucus harvested from primary epithelial nasal cultures grown at air-liquid interface (nasal mucus) or aqueous citric acid/Na_2_HPO_4_ buffer. Figure 1 summarizes the inactivation times (here expressed as the time to reach a 99% infectivity loss) as a function of pH. All viruses were stable in all matrices at neutral pH, with inactivation times of several days. From pH 6 to 4, IAV inactivation times decreased from days to seconds, or by about five orders of magnitude. This decrease was evident in all matrices studied. It is noteworthy that inactivation in nasal mucus, which is most representative of the matrix comprising expiratory aerosol particles, is well described by SLF. However, inactivation times did depend on the SLF concentration. Specifically, we determined IAV inactivation at three different levels of SLF enrichment (1× and 18× SLF, determined experimentally; 24× SLF, determined by extrapolation), corresponding to water activities *a*_*w*_ = 0.994, 0.8 and 0.5. This represents the fluid in equilibrium with a gas phase at 99.4%, 80% and 50% RH, i.e. from physiological equilibrium to common indoor conditions. While inactivation times in aqueous buffer, 1× SLF and nasal mucus were very similar, 18× enrichment of the SLF coincided with an increase in inactivation time by up to a factor 56 (blue triangles in Fig. 1). This protective effect of concentrated SLF was most prominent around the optimal pH for A/WSN/33 viral fusion of ∼ 5.1 (28). Coronaviruses were less affected by acidic pH than IAV. Both, SARS-CoV-2 and HCoV-229E remained largely stable down to pH 3, where their inactivation still required 24 hours. When further decreasing pH down to 2, the inactivation times rapidly reduced to < 10 seconds for SARS-CoV-2, but never dropped below 2 hours for HCoV-229E. Compared to aqueous buffer, SLF provided some protection against inactivation below pH 3, both at 1× and 5× SLF concentrations (while measurements for pH < 3 in 18× SLF were not possible due to precipitation). The measured differences in pH-sensitivities between IAV and the coronaviruses may be explained by their different mechanisms of virus entry into host cells. IAV relies on an acid-induced conformational change in haemagglutinin during endosomal entry. This conformational change is irreversible (29); if IAV encounters the fusion pH (typically pH < 5.5) outside the host cell, e.g. whilst within an aerosol particle, the acid-triggered haemagglutinin can no longer bind to host-cell receptors and the virus is inactivated. Conversely, the spike glycoprotein of coronaviruses becomes fusion competent through cleavage by host proteases, instead of relying on acidic pH triggering conformational changes(30). The different behavior of SARS-CoV-2 and HCoV-229E at pH < 3 remains unclear.

**Fig. 1.**
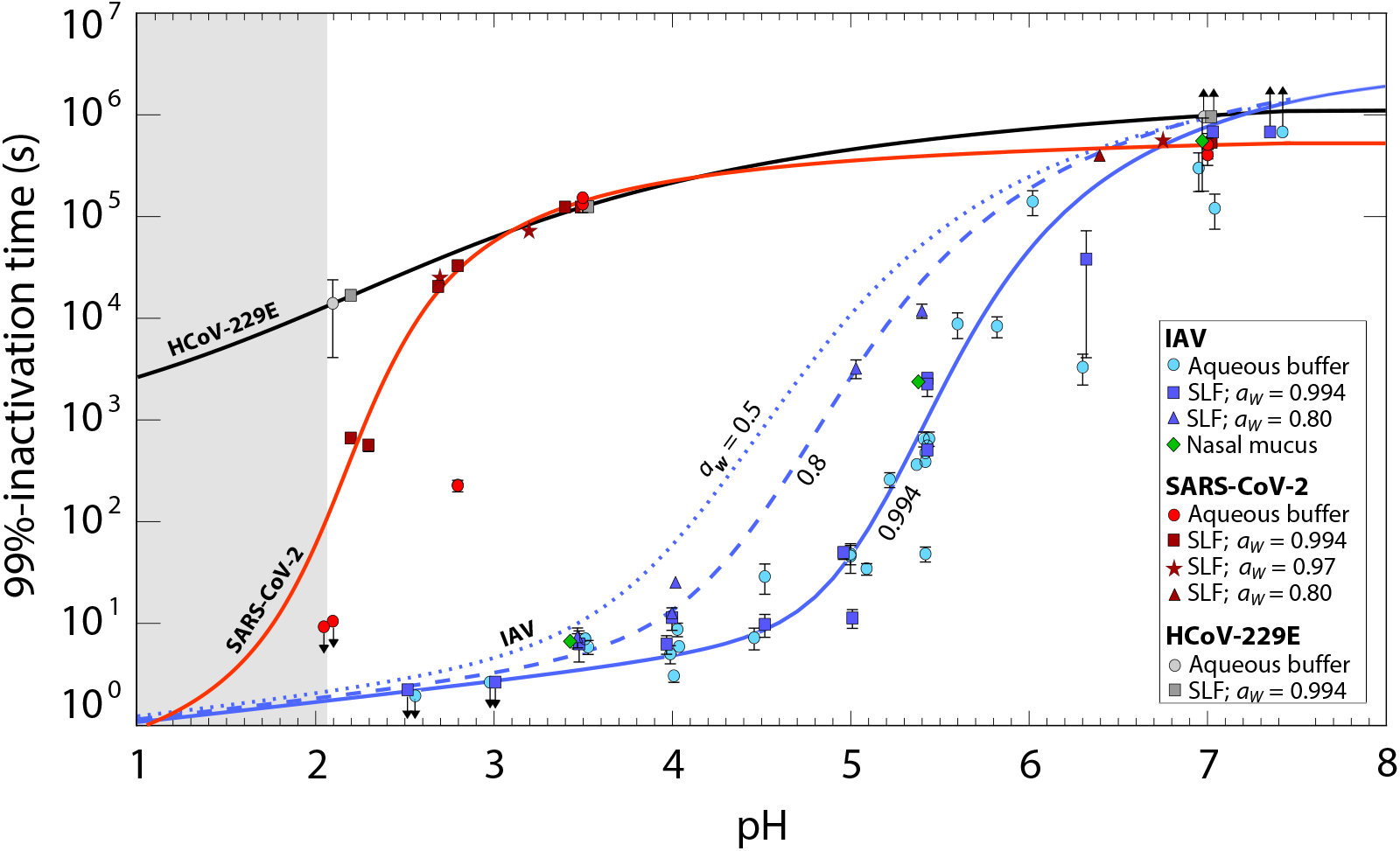
Time required for 99% titer reduction of influenza A virus (IAV), SARS-CoV-2 and human coronavirus HCoV-229E in various bulk media. Data points represent inactivation times in aqueous citric acid/Na_2_HPO_4_ buffer, synthetic lung fluid (SLF) or nasal mucus with pH between 7.4 and 2, measured at 22°C. SLF concentrations correspond to water activity *a*_*w*_ = 0.994 (1× SLF; squares), *a*_*w*_ = 0.97 (5× SLF; stars) and *a*_*w*_ = 0.8 (18× SLF; triangles); buffer (circles) and nasal mucus (diamonds) correspond to *a*_*w*_ ∼ 0.99. Each experimental condition was tested in replicate with error bars indicating 95% confidence intervals. While IAV displays a pronounced reduction in infectivity around pH 5, SARS-CoV-2 develops a similar reduction only close to pH 2, and HCoV-229E is largely pH-insensitive. Solid lines are arctan fits to SLF data with *a*_*w*_ = 0.994 (blue: IAV; red: SARS-CoV-2; black: HCoV-229E). The dashed line is an arctan fit to the SLF data with *a*_*w*_ = 0.80. The dotted line is an extrapolation to *a*_*w*_ = 0.5 (24× SLF). Upward arrows indicate insignificant change in titer over the course of the experiment, and downward arrows indicate inactivation below the level of detection at all measured times. The fitted curves below pH 2 (grey shaded aera) are extrapolated with high uncertainty. Examples of measured inactivation curves are shown in Fig. S1. The arctan fit equations, which are also used for the model simulations, are given by Eqns. S21, S22, and S23.

### Thermodynamics and diffusion kinetics of expiratory particles

While Figure 1 shows the pH that must be attained in the aerosol particles for rapid virus inactivation, it lacks information on aerosol particle pH after exhalation into indoor air. To model the pH in these particles it is essential to know the particle composition in thermodynamic equilibrium (liquid water content), as well as the kinetics that determine how rapidly the equilibrium is approached (water and ion diffusion coefficients). To obtain this information, we measured thermodynamic (equilibrium) and kinetic (diffusion-controlled) properties of individual micrometer-sized SLF and nasal mucus particles levitated contact-free in an electrodynamic balance (EDB). Each particle was exposed to prescribed changes in RH (see Fig. 2).

**Fig. 2.**
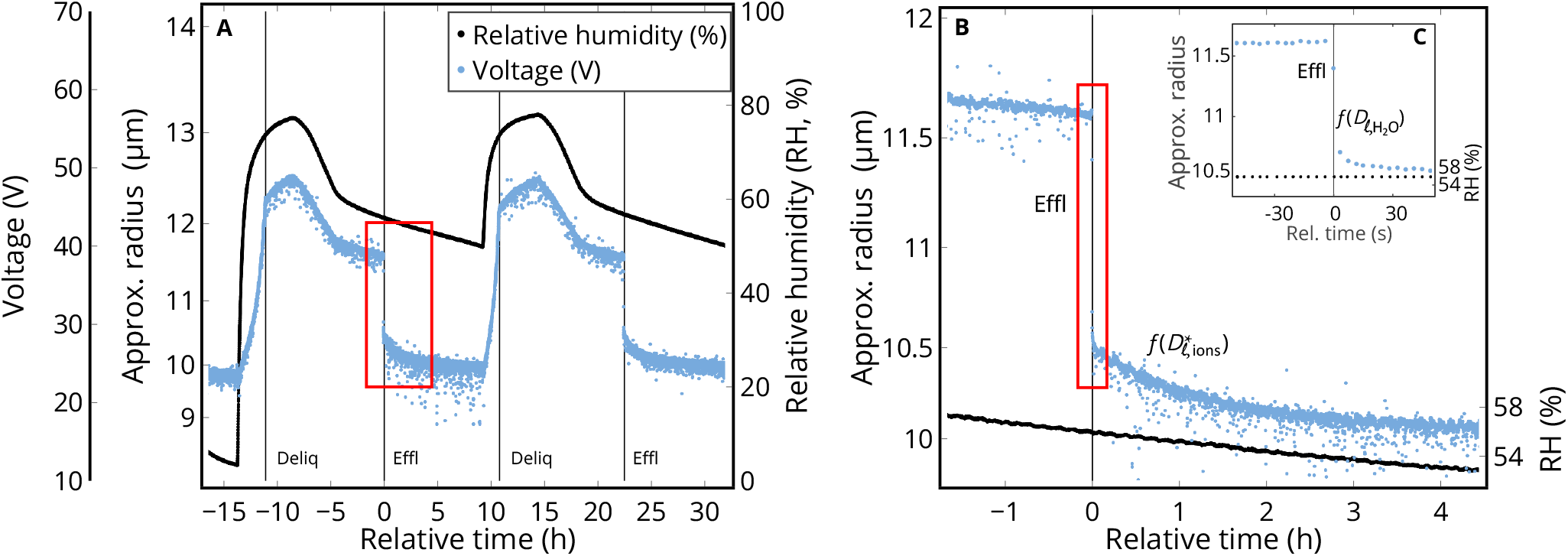
Measured hygroscopicity cycles of a synthetic lung fluid (SLF) particle in an electrodynamic balance (EDB) forced by prescribed changes in relative humidity (RH). The voltage required to balance the particle in the EDB against gravitational settling and aerodynamic forces is a measure of the particle’s mass-to-charge ratio, allowing the particle radius *R* to be estimated. (*A*) Two humidification cycles of an SLF particle with a dry radius *R*_0_ ∼ 9.7 µm. The experiment spanned about 2 days with slow humidity changes, allowing the thermodynamic and kinetic properties of SLF to be determined. Deliquescence/efflorescence points are marked by “Deliq/Effl”. (*B*) Zoom on the drying phase (red box in (*A*)) with salts in the droplet (mainly NaCl) efflorescing around 56% RH (black line): very fast initial crystal growth (< 10 s) with rapid loss of H_2_O from the particle, followed by slow further crystal growth (1 h). The latter is caused by the abrupt switch from H_2_O diffusion to the diffusion of Na^+^ and Cl^−^ ions through the viscous liquid, resulting in an ion diffusion coefficient of 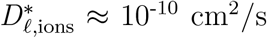. The insert (*C*) highlights the minute before and after efflorescence, which allows a lower bound of the H_2_O diffusivity to be determined, namely 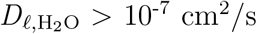.

Figure 2*A* shows two moistening/drying cycles of an SLF particle obtained over a period of two days. They allow determination of the particle equilibrium composition (water content or mass fraction of solutes, see Fig. S2*A*) during time intervals with slowly changing RH. The particle clearly takes up and loses water when the RH is changed. It has a size growth factor at 90% RH of 1.3 (see also Fig. S3) and deliquesces at 75%, indicating that NaCl is the predominant salt in the particle. Nasal mucus shows a similar size growth, but deliquesces over an RH range of 55 to 70%, indicating that it contains significant amounts of other salts (Fig. S3). We have no evidence for liquid-liquid phase separation in any of these particles (Fig. S4A and S5) but Mie-Resonance spectra indicate inhomogenities in the particles even at high RH.

The kinetics of water uptake/loss as derived from periods with rapid RH change or efflorescence are highlighted in Fig. 2. Figure 2*B* zooms in on one efflorescence event, first showing rapid water loss (< 10 s), then switching to a much slower rate of water loss over the next hour. This two-stage diffusion process was confirmed in measurements of additional SLF and nasal mucus particles (see Fig. S6). We attribute the fast process to an initial dendritic growth of an NaCl crystal (Fig. S4*A-C*), which ends abruptly when the crystal reaches the droplet surface, followed by a slow crystal growth mode (Fig. S4*D*). Initially, crystal growth is limited by the liquid phase diffusivity of water molecules with 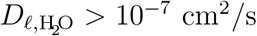 (Fig. 2*C*), which is expelled from the particle as long as water activity is still high. Subsequently, the slow crystal growth is limited by the diffusivities of Na^+^ and Cl^−^ ions through the progressively viscous liquid to the crystal (Fig. S4*D*). From Figs. 2*B* and S4D we estimate the ion diffusion coefficient to be about 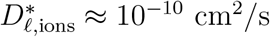, which determines the low rate of continued loss of water molecules. The diffusion coefficients determined in this way are “effective” (indicated by a star), as they represent the molecular diffusivities under the specific morphological conditions associated with the dendritic growth of the salt crystals inside the droplets (see next section for details on how these diffusion coefficients were further constrained).

Independent of the exact thermodynamic equilibrium state of the particles, our results demonstrate that SLF as well as nasal mucus show a clear diffusion limitation for ions. In contrast, water diffusion in SLF and nasal mucus remains fast even when RH is low. This continuous, rapid diffusion of water indicates that SLF and nasal mucus do not form diffusion-inhibiting, semisolid phase states such as those recently reported by others in particles containing model respiratory compounds (31).

### Biophysical model of inactivation in expiratory aerosol particles

Only the combination of the virological bulk phase data (Fig. 1) with the microphysical aerosol thermodynamics (vapor pressures and activity coefficients) and kinetics (Figs. 2 and S7) allows the pH attained in the aerosol particles and the resulting rates of viral inactivation, to be determined. Thus, the virological and microphysical data were combined as input for a multi-layer Respiratory Aerosol Model (ResAM). ResAM is a biophysical model that simulates the composition and pH changes inside an expiratory particle during exhalation, and determines the impact of these changes on virus infectivity (see section “Biophysical modeling” and Supplementary Material). The model performs calculations for particles of selectable size (from 20 nm to 1 mm) with a liquid phase composed of organic and inorganic species representative of human respiratory fluids S1 (more detail in). It takes account of diffusion in the gaseous and condensed phase, vapor pressures, heat transfer, deliquescence, efflorescence, species dissociation, and activity coefficients due to electrolytic ion interactions (see Tables S2, S3). Ultimately, ResAM computes the species distribution and their activity in the liquid, the resulting pH, and the corresponding virus inactivation rates as function of time and of the radial coordinate within the particle.

When RH changes are slow, the measured mass fraction of solutes in SLF as a function of RH allows the model thermodynamics to be constrained (Fig. S2*B*). Under thermodynamic equilibrium conditions the model captures the mass fraction of solutes along the deliquesced and effloresced branches of the particle reasonably well. However, only after kinetic effects (ion and water diffusivities) are also taken into account does the model accurately reproduce the solute composition curve along the deliquesced branch. This demonstrates that even when RH changes are slow (raising RH from 50% to 70% in over one hour), kinetics cannot be neglected.

For rapidly evaporating expiratory particles, kinetics effects are even more critical. By matching the model to the fast changes during the efflorescence and deliquescence processes, ion diffusion coefficients can be derived for different water activities. Interpolation together with literature data in dilute conditions yields 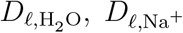, and 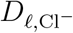 (for details see Fig. S7*D*). Other neutral species, cations and anions are treated accordingly, scaled with their infinite dilution values (see Supplementary Material).

As an example, Fig. 3 shows the evolution of the physicochemical conditions within an expiratory particle with 1 µm initial radius during transition from nasal to typical indoor air conditions with 50% RH (Table S1), and the concomitant inactivation of IAV and SARS-CoV-2 contained within the particle. The rapid loss of water leads to concentration of the organics and salts, to the point when NaCl effloresces. Nitric acid from the indoor air enters the particle readily, lowering its pH from an initial value of 6.6 (resulting from the high concentrations of CO_2_ and NH_3_ in the exhaled air) to pH 5 within ∼ 10 s. This, in turn, pulls NH_3_ into the particle, partly compensating the acidification. The pH further decreases to ~ 4 within 2 minutes, then slowly approaches pH 3.7 due to further uptake of HNO_3_ from the room air. This result confirms the importance of trace gases in determining the pH of indoor aerosol particles (25). If only CO_2_ is considered, its volatilization from the particle would lead to an expected increase in pH after exhalation (32). Owing to aerosol acidification, rapid influenza virus inactivation occurs at ∼2 minutes, whereas SARS-CoV-2 (and the even more pH-tolerant HCoV-229E) remain infectious.

**Fig. 3.**
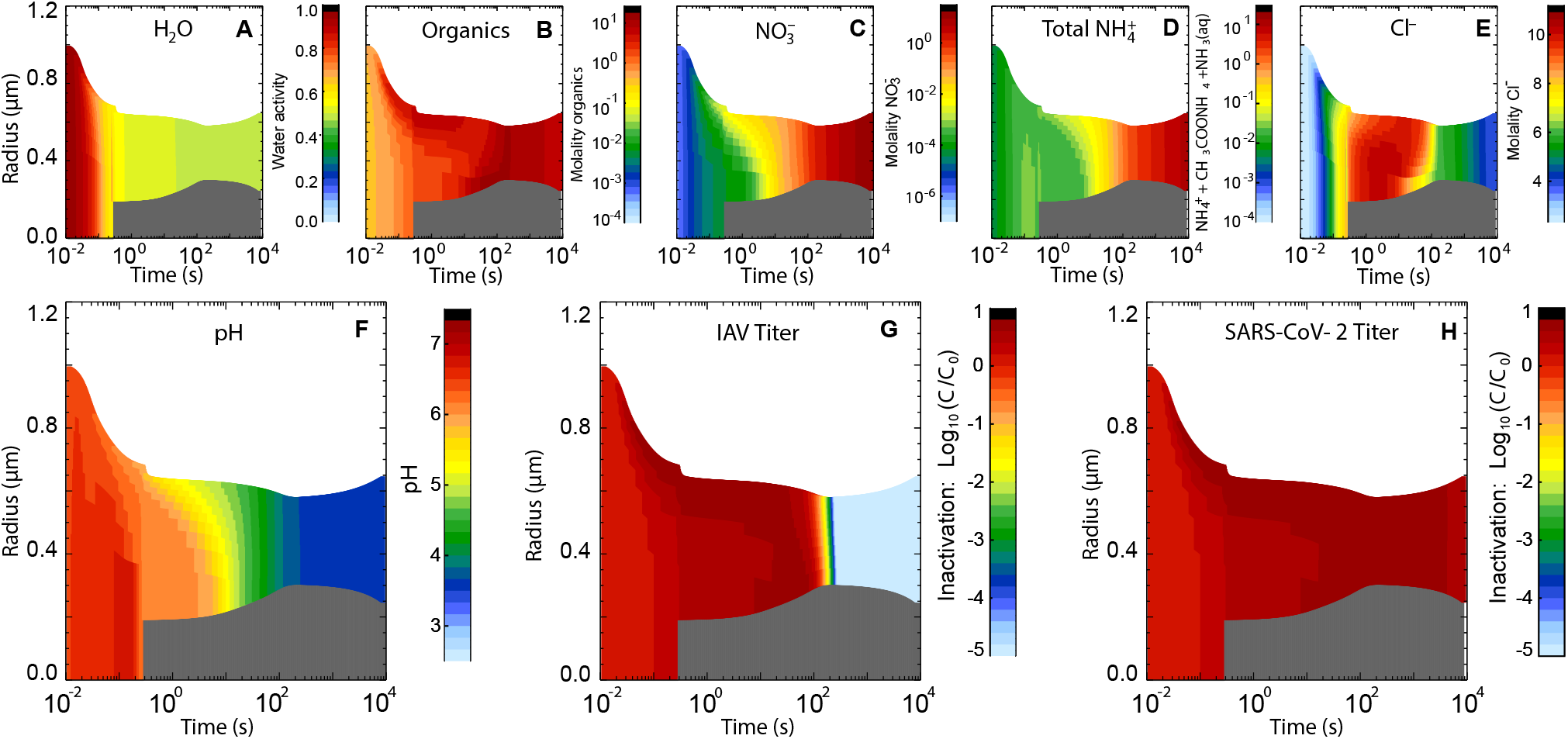
Evolution of physicochemical conditions within a respiratory particle leading to inactivation of trapped viruses during the transition from nasal to typical indoor air conditions, modeled with ResAM. The initial radius of the particle is 1 µm. Thermodynamic and kinetic properties are those of synthetic lung fluid (SLF, see Fig. 2 and Table S1). The indoor air conditions are set at 20°C and 50% RH (see Fig. S8 for the corresponding depiction of physicochemical conditions at 80% RH). The exhaled air is assumed to mix into the indoor air using a turbulent eddy diffusion coefficient of 50 cm^2^/s (see (33) and Supplementary Material, section “Mixing of the exhaled aerosol with indoor air”). The temporal evolution of gas phase mixing ratios is shown in Fig. S20. The gas phase compositions of exhaled and typical indoor air are given in Table S4. Within 0.3 s, the particle shrinks to 0.7 µm due to rapid H_2_O loss, causing NaCl to effloresce (grey core). The particle then reaches 0.6 µm within 2 minutes due to further crystal growth, after which it slowly grows again due to coupled HNO_3_ and NH_3_ uptake and HCl loss. ResAM models the physicochemical changes in particles including (*A*) water activity, (*B*) molality of organics, (*C*) NO^**-**^_**3**_ (resulting from the deprotonation of HNO_3_), (*D*) molality of total ammonium, (*E*) molality of Cl^-^, (*F*) pH, as well as inactivation of (*G*) IAV and (*H*) SARS-CoV-2 (decadal logarithm of virus titer *C* at time *t* relative to initial virus titer *C* _0_).

Inactivation times vary with particle size: larger droplets take longer to reach low pH than smaller ones as they are impeded by longer diffusion paths of the relevant molecules (mainly HNO_3_ and NH_3_) or ions through both the air and liquid phases. The black line in Fig. 4*D* illustrates this relationship for IAV, showing 99% inactivation after about 2 minutes in particles with radii < 1 µm, but longer than 5 days for millimeter-sized particles. As a rule of thumb, a 10-fold increase in particle size leads to roughly a 10-fold increase in IAV inactivation time under typical indoor conditions. Conversely, the black line in Fig. 4*E* for SARS-CoV-2 shows that inactivation is inefficient for SARS-CoV-2, irrespective of particle size.

**Fig. 4.**
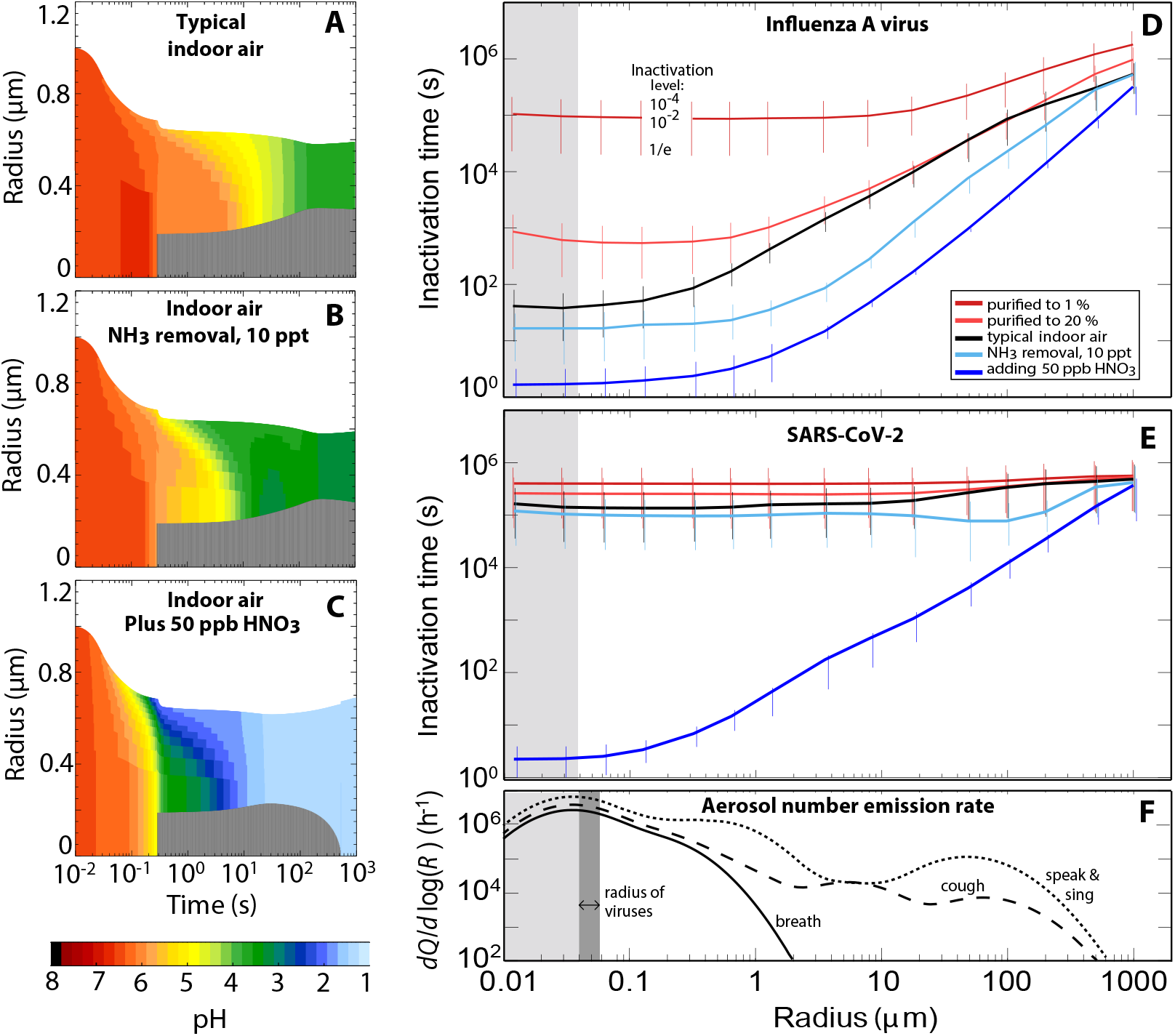
Impact of airborne acidity on virus inactivation in expiratory particles. (*A*) Modeled pH value in a particle with properties of synthetic lung fluid with initially 1 µm radius exhaled into air (20°C, 50% RH) with typical indoor composition (same as Fig. 3*F*). (*B*) Same as (*A*), but for indoor air with NH_3_ reduced to 10 ppt, e.g., by means of an NH_3_ scrubber, reducing the time to reach pH 4 from 2 minutes to less than 10 seconds. (*C*) Same as (*A*), but in indoor air enriched with 50 ppb HNO_3_, reducing the time to reach pH 4 from 2 minutes to less than 0.5 seconds. (*D* and *E*) Inactivation times of IAV and SARS-CoV-2 as function of particle radius under various conditions: indoor air with typical composition (black), depleted in NH_3_ to 10 ppt (light blue), enriched with 50 ppb HNO_3_ (dark blue), or purified air with both, HNO_3_ and NH_3_, reduced to 20% or 1% (red). Whiskers show reductions of virus load to 10^−4^ (upper end), 10^−2^ (intersection with line) and 1/e (lower end). The exhaled air mixes with the indoor air by turbulent eddy diffusion (same as Fig. 3); for sensitivity tests on eddy diffusivity see Figs. S11*B* and S12*B*. The gas phase compositions of exhaled air and the various cases of indoor air shown here are defined in Table S4. (*F*) Mean size distribution (36) of number emission rates of expiratory aerosol particles (*dQ/d*log(*R*)) for breathing (solid line), speaking and singing (dotted line) and coughing (dashed line). Dark grey range indicates virus radii. Light grey shading shows conditions for particles smaller than a virus, referring to an equivalent coating volume with inactivation times indicated. (Radius values in (*D*)-(*F*) refer to the particle size 1 s after exhalation.)

Inactivation times for both IAV and SARS-CoV-2 can be greatly reduced if the indoor air is slightly acidified. This can be achieved by either removing basic gases or adding acidic ones, provided that the gaseous acid molecules meet two conditions: their volatility must be sufficiently low, such that they readily partition from the gas phase to the condensed phase, and, once dissolved, they must be sufficiently strong acids to overcome any pH buffering by the particle matrix. Figure 4 compares the aerosol pH in typical indoor air (panel *A*) with that in air depleted in NH_3_ to 10 ppt (panel *B*), or enriched with 50 ppb HNO_3_ (panel *C*). This concentration of HNO_3_ is well below legal 8-h exposure thresholds (0.5 ppm (34) or 2 ppm (35)).

Scrubbing of NH_3_ reduces the time to reach an aerosol pH of 4 from minutes to seconds. Correspondingly, IAV inactivation times decrease by up to an order of magnitude (light blue lines in Fig. 4*D-E*). This acceleration is mostly limited to particles in the 2 to 5 µm size range, which are minor contributors to the exhaled aerosol (Fig. 4*F*). Furthermore, NH_3_ scrubbing does not affect SARS-CoV-2 inactivation, because the aerosol pH remains in this virus’ stability range (Fig. 1).

A much stronger effect is observed for the addition HNO_3_. Here, 50 ppb allows the aerosol pH value to drop below 2, which is required for efficient SARS-CoV-2 inactivation (Fig. 1). For comparison, enriching the air with the more volatile and weaker acetic acid at concentrations below exposure threshold values could not achieve this, see Fig. S9. The dark blue lines in Fig. 4*D-E* show the resulting inactivation times for IAV and SARS-CoV-2 (and Fig. S10 for HCoV-229E) as a function of particle radius. Remarkably, inactivation times of SARS-CoV-2 diminished by 4-5 orders of magnitude compared to typical indoor air (black lines). For particles with radii < 1 µm, which constitutes the majority of expiratory particles (see panel *F*), inactivation is expected to occur within 30 s.

While an enrichment of acidic gases in air leads to an acceleration of IAV and SARS-CoV-2 inactivation, the depletion of these gases, for instance by air filtration, has the opposite effect. It is well-known that concentrations of strong inorganic acids, such as HNO_3_, are lower indoors than outdoors by at least a factor 2, and in buildings with special air purification, such as museums and libraries, by factors 10-80 (25). If air is purified to contain only a fraction of the initial trace gas concentrations (see Table S4), the aerosol pH increases compared to typical indoor air and intermittently reaches neutral or even slightly alkaline values (up to pH 8.4 in particles with 5 µm radius in air purified to 1%). As a result, air purification is expected to enhance virus persistence, especially for IAV, as indicated by the red curves in Fig. 4*D-E*.

To validate the model results, we compared published inactivation data for aerosolized IAV and SARS-CoV-2 obtained in rotating drum experiments with inactivation times estimated by ResAM (Figures S13 and S14 and Supplementary Text). For both viruses, modelled and measured inactivation times exhibited similar trends as a function of RH. For IAV, measured inactivation times are consistent with ResAM predictions for experiments conducted in partly purified air, as is expected for rotating drum experiments. The comparison with SARS-CoV-2 is inconclusive, because of the wide scatter in the experimental data. However, ResAM predictions fall within the range of measured inactivation times. Given the importance of semi-volatile acids and bases for inactivation, further model validation should include inactivation times measured in aerosol experiments under well-known air compositions, including the presence of HNO_3_.

### Management of airborne transmission risks

Given the high pH sensitivity of many viruses (18, 37–39) and the readiness of expiratory aerosol particles for acidification, we next investigated the extent to which the modification of indoor air composition could mitigate the risk of virus transmission. To this end, we consider a ventilated room with occupants who exhale aerosol containing infectious viruses. We further make the assumption that, given the low concentration of airborne viruses, the transmission risk is directly proportional to the infectious virus concentration, respectively inhalation dose. We use the term “relative risk of transmission” to express how the risk changes from standard conditions (here typical indoor air according to Table S4) compared to air slightly enriched by HNO_3_, scrubbed of NH_3_, or air that has been purified.

For the ventilated room we assume steady-state conditions where the exhalation defines the source of virus, which is balanced by three sinks, namely air exchange through ventilation, aerosol deposition, and pH-moderated virus inactivation within the aerosol particles (see Supplementary Material). We describe the virus source by the mean size distributions of number emission rates of expiratory aerosol particles (Fig. 4*F*) and assume each particle with radius > 50 nm to carry one virus irrespective of size. We describe the virus sinks by expressing ventilation by Air Change per Hour (ACH, mixing ventilation), applying mean aerosol deposition rates (40), and computing the inactivation rates similar to Fig. 4*D-E*. This allows the airborne viral load and, thus, the relative risk of transmission, to be calculated as displayed in Fig. 5 for IAV and SARS-CoV-2 (and Fig. S15 for HCoV-229E). Black bars show the results for typical indoor conditions, light blue bars indicate air from which NH_3_ was scrubbed to 10 ppt, dark blue bars an enrichment of HNO_3_ to 50 ppb, and red bars indicate purification of air to 20% or 1% of trace gases (see Table S4).

**Fig. 5.**
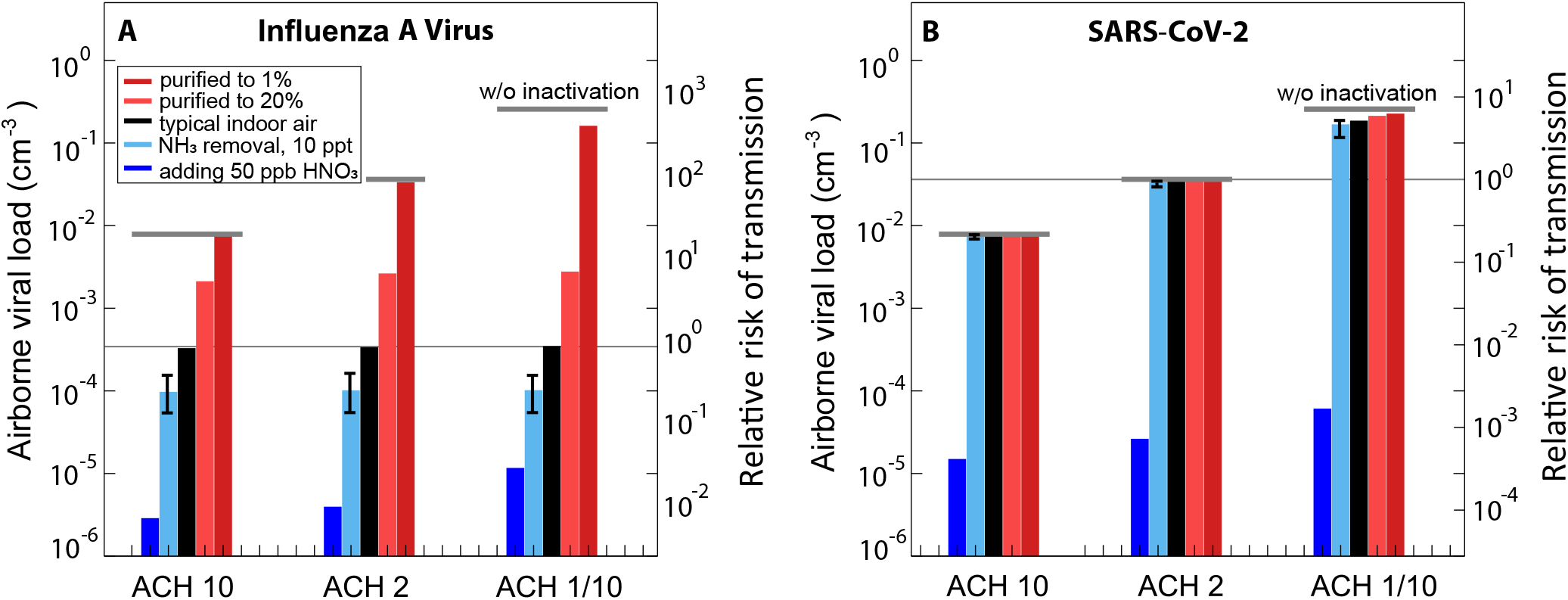
Airborne viral load and relative risk of IAV and SARS-CoV-2 transmission under different air treatment scenarios. The airborne viral load (# viruses per volume of air) and relative risk of transmission of (*A*) IAV and (*B*) SARS-CoV-2 is calculated for a room with different ventilation rates, Air Changes per Hour (ACH), and subject to various air treatments, such as NH_3_ removal, addition of HNO_3_, or supply of purified air. The room is assumed to accommodate one infected person per 10 m^3^ of air volume, emitting virus-laden aerosol by normal breathing (solid curve in Fig. 4*E*), and assuming one infectious virus per aerosol particle. (Corresponding plots assuming a virus concentration that is proportional to the size of the aerosol particles are shown in Fig. S16.) Steady state virus loading, i.e. number of infectious viruses per cubic centimeter of air (left axes), is calculated as the balance of exhaled viruses and their removal by ventilation, deposition, and inactivation. Results are shown for three different ventilation strengths. Virus inactivation is calculated according to Fig. 4*D-E*, starting from radius 0.05 µm. Note, that the mixing speed of the exhalation plume with indoor air depends on ACH (following (33), see Supplementary Text). Right axes show the transmission risk under these treatments relative to the risk in a room with typical indoor air (see Table S4) and ACH 2 (thin horizontal line). Typical indoor air is shown by black bars, filtered air with removal of trace gases to 20% and 1% by red bars, air with NH_3_ removed to 10 ppt by light blue bars, and air enriched with 50 ppb HNO_3_ by dark blue bars. The lower and upper limits for the case with NH_3_ removal shows the range of possible HNO_3_ release from the background aerosol particles after removing NH_3_ from the indoor air (see Table S4). Thick grey horizontal lines indicate the viral load and relative transmission risk in the absence of any inactivation. Results for 2 and 5 ppb HNO_3_ are shown in Fig. S17. Results for HCoV-229E, along with analogous analyses for coughing and speaking/singing are shown in Fig. S15.

The results are unambiguous: adding 50 ppb HNO_3_ diminishes the relative risk of transmission of IAV by a factor of ∼20 and of SARS-CoV-2 by a factor of 800 in rooms with ACH 2. Interestingly, HNO_3_ addition outperforms an increase in ventilation from ACH 2 to ACH 10, which for SARS-CoV-2 leads to a mere dilution by a factor 5 and for IAV does not help at all (black bars in Fig. 5). This is because in typical indoor air, the 99%-inactivation times of IAV are much shorter than the air exchange times, such that virus dilution plays no role. Upon enrichment of the air with 50 ppb HNO_3_, inactivation times of IAV and SARS-CoV-2 drop to only a few seconds for small particles (Fig. 4*D-E*) and are now on the same time scale as HNO3 transport through the gas phase to the droplet by ACH-induced eddy diffusion (see Supplementary Text). In this scenario, higher ACH leads to a faster mixing of the HNO_3_-enriched air into the exhaled plume, resulting in faster acidification of the exhaled aerosol, and hence a lower relative risk of transmission at higher ACH (dark blue bars in Fig. 5). In contrast, adding 50 ppb HNO_3_ only has a moderate impact on HCoV-229E (Figs. S10 and S15).

In comparison, NH_3_ scrubbing has a small, though noticeable effect on the relative risk of IAV transmission, which is reduced by a factor 2-3 depending on the human activity (Figs. S15 and S16). This approach, however, is ineffective for SARS-CoV-2 or HCoV-229E, highlighting the importance of ventilation in such a scenario.

Finally, the ResAM estimates for purified air with significant reduction of trace gases (red bars) are also striking. While even normal air conditioning systems with air filters can lead to a reduction in “sticky” molecules such as HNO_3_ (41), acid removal is likely even more pronounced in museums, libraries or hospitals with activated carbon filters (25). In such public buildings, the relative risk of IAV transmission can increase significantly compared to buildings supplied with unfiltered outside air.

In summary, we demonstrate that the control of aerosol pH is a critical tool in the mitigation of airborne virus transmission. A significant abatement in transmission risk can be achieved by air acidification. For strongly pH-sensitive viruses (e.g., IAV), mere scrubbing of NH_3_ from indoor air suffices to bring about a modest reduction in the airborne viral load. A greater effect that also extends to more acid-tolerant viruses (e.g., SARS-CoV-2) results from air enrichment with an acidic gas. Here we evaluated the use of HNO_3_ for this purpose, though alternative acids may achieve similar results. An effective reduction in viral load can already be achieved by applying HNO_3_ at levels lower than 10% of the legal exposure thresholds (34, 35). We therefore expect that the resulting acid exposure will not cause harmful effects on human health. Nevertheless, future studies should investigate the consequences of acid accumulation in indoor air on the microbiome and immune response in the respiratory tract. Additionally, methods are needed for real-time monitoring of aerosol pH, both to prevent acid overexposure and to ensure efficient virus inactivation. Despite the current unknowns, targeted regulation of aerosol pH promises profound positive effects on airborne virus control. Practices that help acidify exhaled aerosols should thus be considered as a strategy to mitigate virus transmission and disease - alongside interventions such as ventilation that mechanically reduce the concentration of airborne viruses (i.e., dilution) and ensure the resupply of acid molecules from outside air.

## Materials and Methods

### Virus propagation, purification and enumeration

Influenza virus strain A/WSN/33 (H1N1) was propagated in Madin-Darby Canine Kidney (MDCK) cells (ThermoFisher) and the coronavirus strain HCoV-229E-Ren (kindly provided by Volker Thiel, University of Bern)(42) was propagated in Huh-7cells (a kind gift from Mirco Schmolke, University of Geneva). SARS-CoV-2 strain BetaCoV/Germany/BavPat1/2020 was obtained from the European Virus Archive GLOBAL (EVA-GLOBAL; Ref-SKU: 026V-03883)(43) and propagated in VeroE6 cells (kindly provided by Volker Thiel, University of Bern). All work with infectious SARS-CoV-2 was performed in an approved biosafety level 3 (BSL3) facility by trained personnel at the Institute of Medical Virology, University of Zurich. All procedures and protective measures were thoroughly risk assessed prior to starting the project and were approved by the Swiss Federal Office of Public Health (Ecogen number A202808/3). The cells are maintained in Dulbecco’s modified Eagle’s medium (DMEM; Gibco) supplemented with 10% Fetal Bovine Serum (FBS; Gibco) and 1% Penicillin-Streptomycin 10’000 U/ml (P/S; Gibco). MDCK, VeroE6 or Huh-7 cell cultures were inoculated with IAV, SARS-CoV-2 or HCoV-229E at a multiplicity of infection of 0.001, 0.001 and 0.01, respectively, for 72 h. Culture supernatants were clarified by centrifugation at 2,500 × g for 10 min, and IAV and HCoV-229E were pelleted through a 30% sucrose cushion at 112,400 × g in a SW32Ti rotor (Beckman) or a AH-629 rotor (ThermoFisher Scientific) for 90 minutes at 4°C. Pellets were recovered in phosphate-buffered saline (PBS) overnight. SARS-CoV-2 stocks were concentrated using Amicon Ultra-15, PLHK Ultracel-PL Membran, 100 kDa tubes (Millipore). The quantification of IAV, SARS-CoV-2 and HCoV-229E titers were done by standard plaque assay on MDCK, VeroE6 or Huh-7 cells respectively with an assay limit of detection (LoD) of 10 PFU/ml. A live-cell Renilla luciferase assay was used as an alternative quantification method to determine HCoV-229E titers in a high-throughput manner. The assay was performed as previously reported (44), on Huh-7 cells at 33°C using 6 µM of the Renilla luciferase substrate EnduRen (Promega) and had an LoD of approximately 500 PFU/ml. Relative Light units were measured at regular intervals with the EnVision multilabel plate reader (PerkinElmer) and the areas under the curve (AUC) were determined. The system was calibrated using reference samples with known titers. All calculations were performed in GraphPad Prism 9.232.

### Matrix preparation

Experiments were conducted in aqueous buffer and in two matrices representative of respiratory liquids: synthetic lung fluid (SLF) and nasal mucus. Aqueous buffer was prepared from 0.1 M citric acid (Acros Organics) and 0.2 M disodium phosphate (Fluka), mixed at varying proportions to obtain the targeted pH. The pH of each buffer was verified using a pH meter (Orion™Versa Star Pro™; ThermoFisher Scientific). SLF was prepared as described by Bicer (45), except that immunoglobulin G was omitted (Supp. Table S1). Hank’s Balanced Salt Solution (HBSS) without phenol red, lyophilized albumin from human serum, human transferrin, 1,2-dipalmitoyl-sn-glycero-3-phosphocholine (DPPC), 1,2-dipalmitoyl-sn-glycero-3-phospho-rac-(1-glycerol) ammonium salt (DPPG), cholesterol, L-ascorbic acid, uric acid and glutathione were purchased from Sigma Aldrich. Liquid SLF was freeze-dried according to the method described by Hassoun *et al*. (46). To create SLF solutions of different enrichments (1×, 10× or 18×), SLF powder was resuspended in the corresponding volume of milli-Q water and was then acidified to the desired pH with 10% of 10× citric acid-phosphate buffer. Note that SLF enrichments beyond 18× were not experimentally feasible. Nasal mucus was prepared from Nasal Epithelial Cells (NEpCs) from three donors, a 73-year-old male, a 50-year-old male and a 41-year-old female (Epithelix, Switzerland, # EP51AB). Cells were cultured in airway epithelium basal growth medium (Promocell) containing 10 µM Y-27632 (Tocris) and the according airway growth medium supplement pack (Promocell). The health and differentiation process of NEpCs into air-liquid interface (ALI) cultures were monitored by measuring the transepithelial electrical resistance and performing immunofluorescence as previously described (47). Mucus was harvested every 2 weeks by incubating NEpC ALI cultures on 24-mm PET filter inserts (Sarstedt) with 500 µl of milli-Q water at 37°C for 10 min and collecting the wash. Mucus from the 3 different NEpC donors was combined and stored at -80°C. For inactivation experiments, mucus was thawed and acidified to the desired pH with 10% of 10x citric acid-phosphate buffer.

### Inactivation experiments

Influenza and coronavirus inactivation were measured at room temperature in 2-ml glass vials (G085S-1-H; Infochroma), 500-µl PCR tubes (Sarstedt), or 1.5 ml plastic tubes (Eppendorf), using a matrix volume between 10 µl and 1 ml. No effect of the reaction volumes on the inactivation kinetics was observed. Each experimental condition was tested in triplicate, except for a subset of SARS-CoV-2 experiments, which were performed in duplicate. IAV stock solutions were diluted with UltraPure Distilled Water (ThermoFisher Scientific) to reach an approximate titer of 10^9^ PFU/ml. Virus stocks were then spiked into each test matrix to an initial experimental titer of 10^7^ PFU/ml for IAV and HCoV-229E or 3×10^6^ PFU/ml for SARS-CoV-2. After spiking, vials were mixed for approximately 5 seconds at medium intensity. Samples were taken at regular time intervals and were neutralized by diluting 1:100 in PBS for infection (PBSi; PBS containing 3% of Bovine Serum Albumin solution 10% in DPBS (Sigma-Aldrich), 1% of P/S and 1% of Ca^2+^/Mg^2+^ 100 mM (CaCl_2_·2 H_2_O and MgCl_2_·6 H_2_O, Acros Organics)). The PBSi has a pH of 7.3. In most experiments with pH < 3.5, PBSi was supplemented with 2% of 10× citric acid-phosphate buffer at pH 7. Dilution in PBSi rather than addition of a strong base was chosen for sample neutralization because the latter approach was found to further decrease the virus titer. Neutralized samples were frozen until enumeration. To determine kinetic parameters, all replicate experiments of a given experimental condition were pooled. Inactivation rate constants were determined from least square fits to the log-linear portion of the inactivation curves, assuming pseudo-first order kinetics,

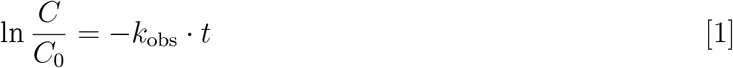

Here, *C* is the virus titer at time *t, C*_0_ is initial virus titer and *k*_*obs*_ is the observed inactivation rate constant. For measurements of *C* below the LoD, *C* was set to the LoD value. 99%-inactivation times (*t*_99_) were determined based on *k*_*obs*_:

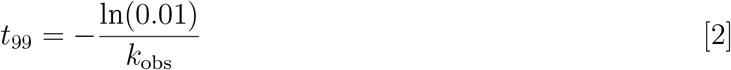

Rate constants and associated 95% confidence intervals were determined using GraphPad Prism v. 9.232. Control experiments were performed to confirm that virus titer loss at low pH could not be attributed to virus aggregation (Fig. S18).

### Measurement of virus aggregate sizes by Dynamic Light Scattering (DLS)

To measure the extent of virus aggregation, IAV was spiked into pH 5 or pH 7 citric acid-phosphate buffer, at a final concentration of 10^10^ PFU/mL. The mean hydrodynamic diameter of viral particles in solution was automatically measured every 2 minutes by DLS using a Zetasizer Nano ZS light scattering instrument (Malvern Panalytical), for a total of 30 minutes. Additionally, samples at pH 5 were neutralized in the cuvette by addition of 2 M Na_2_HPO_4_ at *t* = 30 min, and rapidly mixed by pipetting. The mean hydrodynamic diameter was automatically measured every 2 min for an additional 60 min. Each buffer alone (no virus) was also measured for 10 min each with no particles detected above 10 µm. All data were analyzed using Zetasizer Software 8.01.4906 (Malvern Panalytical).

### SLF characterization by (cryo-)transmission electron microscopy and DLS

Transmission electron microscopy (TEM) was conducted using 15 µl of freshly prepared SLF incubated for 2 minutes on a glow-discharged carbon-coated copper grid (400 mesh). After incubation the grid was washed with solution containing only HBSS (Table S1) and stained with uranyl acetate 2% for 30 seconds. Observations were made using a Tecnai F20 electron microscope (Thermo Fisher, Hillsboro, USA) operated at 200 kV. Digital images were collected using a direct detector camera Falcon III (Thermo Fisher, Hillsboro, USA) 4098 × 4098 pixels using a defocus range between -1.5 µm and -2.5 µm. The characterization of SLF by cryo-transmission electron microscopy (cryo-TEM) was performed using 5 µl of freshly prepared SLF, which was applied onto lacey carbon film grids (300 microMesh, EMS). The grid was blotted in an automatic plunge freezing apparatus (Thermo Fisher, Hillsboro, USA) to control humidity and temperature. Observation was made at -170° C on the Tecnai F20 electron microscope, operated at 200 kV and equipped with a cryo-specimen holder Gatan 626 (Warrendale, PA, USA). Digital images were drift corrected using the camera Falcon III (Thermo Fisher, Hillsboro, USA) 4096 × 4096 pixels. The diameter of the particles in SLF solution was measured by DLS after mixing by vortex for 2 min.

### EDB measurements of aerosol thermodynamics and diffusion kinetics

We used an electrodynamic balance (EDB) to measure the thermodynamic and kinetic properties of micrometer-sized SLF and nasal mucus particles under controlled conditions in the gas phase. The EDB is also called particle “trap”, as it stabilizes (or traps) a slightly charged particle by electric fields in contact-free levitation. Therefore, it enables to investigate droplets, which are highly supersaturated with respect to precipitation of NaCl and other salts. These supersaturated states (see, e.g., efflorescence-deliquescence hysteresis in Fig. S3) occur regularly during exhalation, but are inaccessible for macroscopic measurements, because the contact with bulk containment walls readily leads to precipitation. Thermodynamic properties such as the particle water content in equilibrium with the gas phase are measured under constant or only very slowly changing conditions in the gas phase. In contrast, rapid changes, as those following crystal nucleation and subsequent efflorescence, enable the EDB to also determine the kinetics of fast crystal growth and diffusion processes inside a trapped particle.

The EDB consists of two hyperboloidal endcap electrodes and a central ring electrode with an AC field that stabilizes the particle horizontally, and a charged single particle is held at the null point of the balance by a DC field established across the endcaps (48). As we described previously (49, 50), we use the EDB to measure the relative changes in mass and radius of a single levitated particle caused by changes in RH with very high precision. In brief, using a droplet-on-demand generator a charged particle of SLF (2.5× recipe concentrations) or of freshly thawed nasal mucus was injected into the temperature-regulated (15°C) and RH-controlled gas flow in the EDB and levitated by the adjustable DC electric field. The particle experiences three forces along the symmetry axis of the EDB, which balance each other:

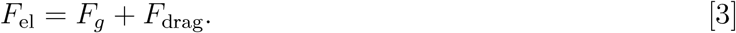

These are the gravitational force and the Stokes drag force

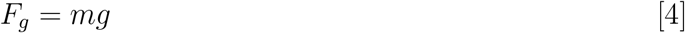

and the Stokes drag force

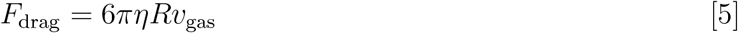

induced by the downward-oriented gas flow. Here, *m* denotes the mass, *g* the gravitational acceleration, *η* the dynamic viscosity of the gas, *R* the particle radius and *v* the velocity of the humidified N_2_ gas flow through the EDB. These two forces are balanced by an electrical force, *F*_el_, required to maintain the particle in the center of the trap,

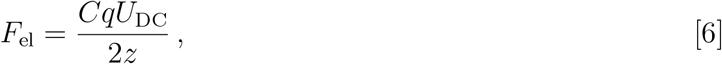

where 2*z* is the minimum distance between the endcap electrodes, *U*_*DC*_ is the DC voltage between the electrodes, *q* is the charge on the particle, and *C* denotes a geometrical constant of the balance. Via Eq. 4, *U*_DC_ depends on the particle mass with a sensitivity to variations caused by loss or uptake of water vapor in the range of 10^*-*13^ to 10^*-*12^ g.

Close to thermodynamic equilibrium, the partitioning of H_2_O between the gas and the condensed phase was examined by slowly (within hours to days) cycling RH in the EDB between dry (< 5%) and humid (ca. 90%) conditions. The total N_2_ flow was set to 20 sccm (controlled by mass flow controllers). The RH was measured with a capacitance sensor, which was calibrated by observing the deliquescence of various salts. Its accuracy is estimated to be ±1.5%.

To calibrate mass changes of trapped particles (or equivalently, their mass fraction of solutes), the particles were exposed to dry conditions (i.e., without H_2_O exchange with the gas phase) to obtain the voltage corresponding solely to the gravitational force and the drag force at various flows. The voltage contributions due to gravity (*U*_0_) and drag (*U*_drag_(0)) were determined from linear regression of the measured voltage (*U*_*m*_) at various flows under dry conditions. Next, the force balance equation 3 was rearranged to obtain:

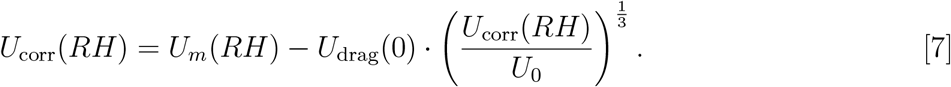

Equation 7 was then analytically solved to retrieve the drag-independent voltage (*U*_corr_(*RH*)). Therefore, at any RH, the mass fraction of solutes is given by *U*_0_*/U*_corr_(*RH*). It was previously shown that certain particle components may induce slow charge loss (51) over a period of hours to days. To correct for that, measurements under similar condition that demonstrated a difference of at least 1 V from cycle to cycle were linearly adjusted prior to the drag force correction.

To complete the analysis of particle composition, the relative change in radius was determined from relative changes in the wavelength of Mie resonances apparent in continuously recorded broad-band backscattering spectra (50, 52, 53) using the Chylek approximation (54). We first estimated the particle radius at 91% RH. Assuming a constant density and refractive index, we then obtained the radii at all other RH (50). This allows a highly accurate determination of particle radii, but we performed this evaluation only on a subset of our measurements, as the method relies on the particles being spherical (which is only approximately true for the inhomogeneous particles of interest in the present work).

### Biophysical modeling

The Respiratory Aerosol Model ResAM is a biophysical model to determine virus inactivation times in the exhalation aerosol as a function of air composition. ResAM is based on a spherical shell diffusion model, which we have previously applied in physical and chemical contexts (55–57). As novel input experimental data, ResAM uses the pH sensitivity of enveloped viruses and of the thermodynamic and kinetic properties of respiratory fluids, both measured in the present work.

ResAM simulates the composition and pH changes inside an exhalation particle during exhalation. Hereby we make the simplifying assumption that the mode of generation (breathing, coughing, singing) does not influences the matrix composition. The model performs calculations for particles of selectable size (from 25 nm to 1 mm) with a liquid composed of H_2_O, H^+^, OH^−^, Na^+^, Cl^−^, CO_2_(aq), HCO_3_^−^, NH_3_(aq), NH_4_^+^, CH_3_COOH(aq), CH_3_COO^−^, CH_3_COONH_4_(aq), NO_3_^−^, as well as two classes of organic compounds with low and a high molecular weight, representative of the lipids and proteins in the lung fluids of interest.

The liquid phase is divided into concentric shells (Fig. S19). The model treats 1-50 shells, depending on particle size (one shell for *r* = 0.02 − 0.1 µm and up to 50 shells for *r* = 1000 µm). The number of shells for each particle stays constant during the exhalation process. The shells are treated in a fully Lagrangian manner, i.e., their thicknesses are calculated from the number of molecules of each species in a shell times their molecular volume, whereby diffusion processes between shells may cause each shell to evolve differently with time.

We take account of vapor pressures 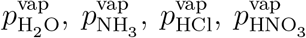 and 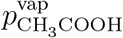 calculated using the Henry’s law coefficients listed in Table S2. The activity coefficients of H^+^, Na^+^, Cl^−^, NO_3_^−^, OH^−^ required for the vapor pressure and concentration simulation are calculated using the Pitzer ion-interaction model(58, 59). The activity coefficients of organic and neutral species are assumed to be unity, i.e., they influence the physicochemical properties of SLF as ideal components simply via Raoult’s Law. SLF contains additional ions in minor concentration (Table S1). For all other minor anions, the activity coefficients of Cl^−^ and the cations the activity coefficients of Na^+^ are used.

We obtain the liquid phase diffusion coefficients of ionic and neutral species as well as the efflorescence RH values from our EDB measurements (Figs. S2, S7). The diffusion coefficients *D*_*ℓ*_ of the involved species at in infinitely diluted water are given in Table S3. The water activity dependence of the diffusion coefficients of H_2_O, Na^+^ and Cl^−^ ions are shown in Fig. S7. In the absence of other information, we assume that *D*_*ℓ*_ of all neutral species have the same dependence on water activity as *D*_*ℓ*,H_2O(*a*_*w*_) scaled with their value at infinite dilution. Similarly, the diffusivities of cations and anions are assumed to have the same as dependence on *a*_*w*_ as Na^+^ and Cl^−^, respectively, again scaled with their dilute solution values from the literature.

When RH decreases below efflorescence RH, we assume the resulting NaCl crystal to reside in the particle center (see Fig. S19). In reality, crystal growth is dendritic (see Fig. S4). We account for the complexities resulting from dendritic crystal growth by using the effective diffusion constants for the crystal growth (see Eqn. S13).

We then use the model to calculate the pH value, and from this the corresponding virus inactivation rates, in each particle shell (see Supplementary Material for details). The gas phase compositions of exhaled air and the indoor air with purification and acidification are shown in Table S4.

Note that the current version of ResAM can readily be further refined beyond the conditions used herein, e.g., to explore the use of alternative acids for aerosol acidification, or to include a greater diversity of respiratory matrices. The two matrices considered in this work - SLF and nasal mucus - have comparable thermodynamic and kinetic properties as well as a similar pH-dependence of viral inactivation. However, we cannot exclude that additional respiratory matrices found in expiratory aerosol plumes (e.g., saliva) exhibit distinct properties (31, 60).

## Data Availability

All data produced in the present work are either contained in the manuscript or will be made available upon acceptance of the manuscript

## Acknowledgements

This work was funded by the Swiss National Science Foundation (grant numbers 189939 and 196729). The authors thank Chuck Haas and Mutian Niu for valuable discussions.

## Author contributions

Conceptualization: AN, SS, TK, TP, UKK, WH

Methodology: AS, BL, IG, LKK, MOP, NB, SCD, SS, TK, TP, UKK

Investigation: AS, BL, IG, KV, LKK, MOP, NB, SCD

Visualization: AS, BL, IG, GM, LKK, KV, SCD, TK, TP

Funding acquisition: AN, SS, TK, TP, UKK, WH

Project administration: TK

Supervision: AN, SS, TK, TP, UKK

Writing–original draft: BL, SS, TK, TP

Writing–review & editing: AN, AS, GM, IG, LKK, MOP, NB, SCD, SS, TP, TK, UKK, WH.

## Data availability

Experimental data and ResAM code will be made available upon manuscript acceptance. A preprint was deposited on medRxiv (doi:10.1101/2022.03.14.22272134). It is made available under a CC-BY-NC-ND 4.0 International license.

