## Supplementary Text, Figures and Tables for "Acidity of expiratory aerosols controls the infectivity of airborne influenza virus and SARS-CoV-2"

**This PDF file includes:**

Materials and Methods
Supplementary Text
Figures S1 to S21
Tables S1 to S4

#### Further ResAM details I: vapor pressures, activity coefficients and pH value

##### Species treated by ResAM

The calculation of composition and pH in the liquid phase of expiratory particles requires knowledge of the vapor pressures of the volatile species and the activities of all species. The particles are highly diluted at the moment of exhalation, but $H_2O$ evaporates rapidly when mixed with dry ambient air. Besides water, the following semi-volatile species, which are important for the pH of exhaled aerosol particles, are considered by ResAM in the exchange with the gas phase: $NH_3$, $HNO_3$, HCl, $CO_2$ and $CH_3COOH$. In the aqueous phase, ResAM treats the following neutral species: $H_2O$, organics (with two representatives, one of proteins and one of lipids, see below), $NH_3$, $CO_2$, $CH_3COOH$ and $NH_4CH_3COO$. Acids, bases, as well as water can dissociate in the aqueous solution, and the degree of dissociation determines the concentrations of ions, and thus the pH value. To this end ResAM treats the cations $H^+$, $Na^+$, ${NH_4}^+$, and the anions $OH^-$, $Cl^-$, ${NO_3}^-$, ${HCO_3}^-$, ${CO_3}^{2-}$, and $CH_3COO^-$. Other minor inorganic species in SLF (see Table S1), such as the anions $Ca^{2+}$, $Mg^{2+}$ and $K^+$, are not explicitly treated, but are subsumed under NaCl with the same activity coefficient as the $Na^+$ ion. This simplification only slightly affects the mass fraction of solutes, the size of the particles, and the computed pH. We group the organic species in SLF into two classes. The first class represents proteins with high molar mass and the second class represents lipids and antioxidants. The mole masses of these two organic classes are determined by fitting ResAM to the EDB measurements (see Figs. 2, S7). An effective mole mass of 1000 Da is obtained for the proteins and 190 Da for lipids and antioxidants, providing the best agreement with the EDB experiments. The organics are treated as ideal components in the fluid (zero activity coefficients), but they affect the physicochemical properties of the fluid via Raoult's law.

##### Aerosol initial conditions

The initial properties of freshly formed exhalation aerosol particles have some influence on their readiness to take up acidic gases from indoor air. In ResAM, we specify the initial conditions for the vapor pressure calculations as follows. The temperature is assumed to be slightly below body temperature (309.1 K) and the water activity of the particle is 0.952. This water activity is the average of water activity of an isotonic solution (Table S1) and the relative humidity at the exit of the nose or mouth (Table S4). Using ResAM, SLF is calculated to have a pH of 6.62 when in equilibrium with 23 550 ppm $CO_2$ and 133 ppb $NH_3$ in the gas phase (i.e., the average of the concentrations of these gases in the exhaled air plume and in typical indoor air; see section "Aerosol initial pH buffering"). For comparison, endoscopic nano-sensor studies of airway lining fluid find pH values *in vivo* of $\sim$ 6.6 in the central airways of healthy humans (61, 62). We use this solution with water activity of 0.952 and pH 6.6 for the initial composition of all simulations in this work (except in Figs. S11A and S12A, which show the sensitivity to other initial pH values). In this reference, the molalities (in mol/kg-$H_2O$) of the involved species are: $H_2O$: 55.51, proteins with an effective molar mass 1000 Da: 0.094, lipids with an effective molar mass 190 Da: 0.322, $Na^+$: 1.371, $Cl^-$: 1.400, other cations: 0.1243, other anions: 0.097, $CO_2$ + ${HCO_3}^-$ + ${CO_3}^{2-}$: $1.908 \times 10^{-3}$, $NH_3$ + ${NH_4}^+$: $1.257 \times 10^{-3}$. Other species are set to be zero.

##### Aerosol initial pH buffering

The pH of SLF is 6.6 in our reference case. However, the concentrations of buffer species such as bicarbonate and ammonia affect pH significantly and have to be treated properly. The acid-base pairs $CO_2(aq)/HCO_3^-$ and $NH_4^+/NH_3(aq)$ are major buffers in respiratory fluids(63, 64). The initial concentrations ($t = 0$) of $CO_2$, $HCO_3^-$ and $CO_3^{2-}$ are calculated assuming equilibrium with 23 550 ppm $CO_2$ in the gas phase (i.e., the mean of the concentrations of $CO_2$ in exhaled air and indoor air, see Table S4), which is an approximation for the internal mixing of inhaled and exhaled air in the respiratory tract. Similarly, we apply a mean concentration of ammonia. Depending on its composition and pH, the respiratory fluid takes up more or less $CO_2$ and, thus, contains more or less carbonate, which acts as an efficient pH buffer. This determines the initial properties of freshly formed expiratory aerosol particles, and also their readiness to subsequently take up acidic gases from the indoor air. The pH $\sim$ 6.6 in the airways of healthy humans can be altered by disease or lung inflammation (62). To estimate the importance of deviations from the reference value of pH 6.6, we performed sensitivity runs with ResAM using an initial pH of 6.2 or 7.0. In the model, these pH values are achieved by adding small amounts of HCl or NaOH to the modeled SLF (while maintaining equilibrium with the mentioned $CO_2$ and $NH_3$ concentrations). In detail, pH 6.2 is achieved by adding 2.63 $\times 10^{-3}$ mol/kg-$H_2O$ of HCl, and pH 7.0 by adding 2.66 $\times 10^{-3}$ mol/kg-$H_2O$ of NaOH (for comparison, the $Na^+$ and $Cl^-$ concentrations in the reference solution are larger than 1 mol/kg-$H_2O$). Consequently, the total concentration of dissolved $CO_2$ (i.e., $CO_2$(aq) + $HCO_3^-$ + $CO_3^{2-}$) varies between $1.1 \times 10^{-3}$ mol/kg-$H_2O$ at pH 6.2 to 3.8 $\times 10^{-3}$ mol/kg-$H_2O$ at pH 7.0. The transition from the release of an aerosol particle inside the respiratory tract to the exits (nose or mouth) is modeled by linear relaxation of the gas phase composition towards exhaled air given by Table S4 (i.e., $CO_2$ ramping up linearly from 23 550 ppm at the moment of particle formation to 46 500 ppm at the nostrils 70 ms later, before relaxing to 600 ppm by eddy mixing with the indoor air). The buffering effect is clear in Figs. S11A and S12A: in the model run with 50 ppb $HNO_3$, an initial pH of 7.0 leads to slower virus inactivation for particles with radii $>$ 1 µm. However, initial pH plays only a minor role for smaller particles. Furthermore, initial pH makes hardly any difference for virus inactivation in typical indoor air (except for IAV in particles $>$ 50 µm).

##### Ion-interaction modeling

The activity coefficients of $H^+$, $Na^+$, $NH_4{}^+$, $Cl^-$, $NO_3{}^-$ and $OH^-$ ions are calculated using the Pitzer ion-interaction model (58, 59, 65). For the anions $CO_3^{2-}$, $HCO_3{}^-$ and $CH_3COO^-$ with minor concentrations, we assume their activity coefficients in SLF to be unity (because of missing interaction parameters). The parameters for NaCl in the Pitzer-ion interaction model were recalculated based on both the data of Chan et al (66) and our own data on pure NaCl shown in Fig. S2. The water activity is calculated considering the contribution of the inorganic and organic species:

$$a_w \approx a_{w,\text{inorganic}} \times a_{w,\text{organic}}, \qquad \text{[S1]}$$

where the water activity of organic species and that of the ions with minor concentrations ($CO_3^{2-}$, $HCO_3^-$, $CH_3COO^-$) are calculated using Raoult's law:

$$a_{w,\text{organic}} = \frac{M_{H_2O}}{M_{H_2O} + \sum_j M_{\text{organics}^j}}, \qquad \text{[S2]}$$

where $M_{H_2O}$ is the molality of water and $M_{\text{organics}^j}$ is the molality of organic species (and $j$ denotes the two organic categories, light and heavy).

##### Dissociation equilibria and pH

The aqueous phase dissociation constants are listed in Table S2. They are used independently in each model shell (following each diffusion time step). The $H^+$ concentration is obtained by setting the net charge in each shell to zero. The pH is then given by

$$\mathrm{pH} = -\log_{10} a_{\mathrm{H}^+}. \qquad \text{[S3]}$$

The vapor pressures of $H_2O$, $NH_3$, $HNO_3$, HCl and and $CH_3COOH$ are then calculated using the Henry's law coefficients listed in Table S2 and the activity coefficients calculated from the Pitzer-ion interaction model.

The difference between partial pressures and vapor pressures of each species determines whether the gas phase is supersaturated or subsaturated with respect to the condensed phase, which in turn decides whether a given species is condensing or evaporating. These pressures (or the corresponding mole fractions) are shown in Fig. S20 for the simulation in Fig. 3. After $\sim 100$ s, the vapor pressure of HCl is higher than its partial pressure, leading to a steady replacement of $Cl^-$ ions by ${NO_3}^+$ ions, and eventually to the full deliquescence of the salt crystal.

#### Further ResAM details II: diffusion processes in expiratory particles

##### *Nernst-Planck Equation and electroneutrality*

By means of our EDB measurements, we identified two distinct stages during crystal growth separated by an abrupt change (see Fig. 2). We interpret this as weak impedance by fast water diffusion in the first stage and strong impedance by slow ion diffusion in the second stage, i.e. $D_{\ell,\mathrm{H_2O}} \gg D_{\ell,\mathrm{ions}}$. However, it is well-known that different ions in a liquid diffuse with different velocities. For example, from isotopic and NMR-measurements in aqueous salt solution, it is known that $D_{\ell,\mathrm{Na}^+} < D_{\ell,\mathrm{Cl}^-} \ll D_{\ell,\mathrm{H}^+}$(67), see Table S3. Given that $H^+$ ions are known to diffuse particularly fast, differences in $Na^+$ and $Cl^-$ diffusivity could influence $H^+$ via electrostatic forces, which has immediate repercussions on particle pH. Therefore, it is critical to understand these microscopic transport processes.

When ions diffuse in a liquid, such as $Na^+$ and $Cl^-$ in an expiratory particle, electroneutrality must be maintained (68), at least on scales much larger than the ions. Otherwise charge-separation would occur, resulting in physically unreasonably high electric fields. Therefore, either an ion of opposite charge diffuses in the same direction (salt diffusion) or an ion of the same charge diffuses in the opposite direction (counter diffusion). In these cases the flux will depend on the individual diffusion coefficients, the concentration gradients and the gradient of the electric potential, which arises from the tendency of one ion to diffuse faster than another. Electroneutrality is violated only on the nano-scale (nano with respect to space and time), as fluctuating hydrodynamics simulations reveal (69). As the $H^+$ ions participate in establishing charge neutrality, this affects the pH, albeit only marginally.

A proper treatment of ion diffusion requires to apply the Nernst-Planck Equation (70). The flux $\vec{j_i}$ of the species $i$ at location $\vec{r}$ inside the particle is given by

$$\vec{j_i}(\vec{r}) = -D_{\ell,i}(\vec{r})c_i(\vec{r})\frac{1}{RT}\nabla\bar{\mu}_i, \qquad \text{[S4]}$$

where $D_{\ell,i}(\vec{r})$ is the liquid-phase diffusion coefficient of species $i$ and $c_i(\vec{r})$ its concentration, $T$ is the temperature of the liquid, and $R$ the universal gas constant. The chemical potential of $i$ in an electric field $\bar{\mu}_i$ is given by:

$$\bar{\mu}_i = \mu_i + z_iF\Phi(\vec{r}) = \mu_0 + RT\ln a_i(\vec{r}) + z_iF\Phi(\vec{r}), \qquad \text{[S5]}$$

where $F$ is the Faraday constant, $z_i$ is the electric charge of species $i$ ($=$ for neutral molecules, $< 0$ for anions, $> 0$ for cations), $a_i(\vec{r})$ its activity, and $\Phi(\vec{r})$ the electric potential. Inserting Eq. S5 into Eq. S4, we obtain the Nernst-Planck equation:

$$\vec{j}_i(\vec{r}) = -c_i(\vec{r}) \times \left( D_{\ell,i}(\vec{r}) \nabla \ln a_i(\vec{r}) + \frac{z_i F}{RT} \vec{E}(\vec{r}) \right), \qquad \text{[S6]}$$

where $\vec{E}(\vec{r})$ is the electric field. Together with the continuity equation, i.e. $\partial c_i / \partial t = -\nabla \cdot \vec{j}_i$, this becomes the diffusion equation for species $i$ in an electric field:

$$\frac{\partial c_i}{\partial t}(\vec{r}) = -\nabla \cdot \vec{j}_i(r) = \nabla \cdot \left( D_{\ell,i}(\vec{r}) c_i(\vec{r}) \nabla \ln a_i(\vec{r}) + c_i(\vec{r}) \frac{z_i F}{RT} \vec{E}(\vec{r}) \right). \qquad \text{[S7]}$$

For neutral species, $z_i$ equals zero, and the Nernst-Planck equation reduces to Fick's first law of diffusion. In Equation S6, $\vec{E}(\vec{r})$ is unknown, but can be obtained from the constraint of charge neutrality in the aerosol, i.e. by demanding zero net charge flux:

$$\sum_i z_i \times \vec{j}_i = \sum_i \left( -z_i \;\; c_i \; D_{\ell,i} \; \nabla \ln a_i - c_i \frac{z_i^2 F}{RT} \vec{E} \right) = 0 \quad , \qquad \text{[S8]}$$

where the sum extends over all species $i$. Therefore:

$$\vec{E} = -\frac{\sum_i z_i \;\; c_i \; D_{\ell,i} \; \nabla \ln a_i}{\sum_i c_i \frac{z_i^2 F}{RT}} \qquad \text{[S9]}$$

Inserting Eq. S9 into Eq. S6 enables us to determine the diffusion flux in the particle. ResAM divides the aerosol particle into shells (Fig. S19) and assumes the exhaled particle to have spherical symmetry, i.e,. net diffusion occurs only along the radial coordinate $r$. Hence, the rate of concentration change $dc_i/dt$ in the shell with radius between $r$ and $r + \Delta r$ equals:

$$\frac{dc_i}{dt} = \frac{4\pi r^2 j_i(r) - 4\pi (r+\Delta r)^2 j_i(r+\Delta r)}{V_i} = \frac{3r^2 j_i(r) - 3(r+\Delta r)^2 j_i(r+\Delta r)}{(r+\Delta r)^3 - r^3} \qquad \text{[S10]}$$

This finalizes the treatment of diffusive fluxes of charged ions and neutral molecules in the interior of aerosol particle by the Nernst-Planck equation. These fluxes need to satisfy the boundary conditions at the interface to the gas phase and to the salt crystal in the case of efflorescence, discussed below.

##### *Boundary condition at the liquid-gas interface*

The flux $\vec{j}_i(r \to r_g)$ in the liquid at the particle interface with the gas phase, $r_g$ ($= r_{n+1}$ in Fig. S19), is given by Eq. S11. This flux needs to consider the exchange of the volatile species with the gas phase, i.e. it needs to equal the flux at $r_g$ in the gas phase. The exchange with the gas phase is calculated using the gas phase diffusion onto a spherical particle surface with radius $r_g$ in steady state, given by(71):

$$j_{\mathrm{surf},i} = 4\pi r_g D_{g,i} \frac{p_i^{\mathrm{vap}}(T_{\mathrm{ptcl}}) - p_i}{RT_{\mathrm{air}}} \qquad \text{[S11]}$$

with $p_i$ being the partial pressure and $p_i^{vap}$ the vapor pressure of species $i$, $D_{g,i}$ is the gas-phase diffusion coefficient of species $i$, $T_{\mathrm{air}}$ the indoor air temperature, and $T_{\mathrm{ptcl}}$ the temperature of the particle.

##### *Boundary condition at the liquid-solid interface and dendritic crystal growth habit*

SLF particles effloresce readily when RH drops below $\sim 56\%$ (Fig. 2). Before we treat the boundary condition for the Nernst-Planck equation, we pay attention to the complex dendritic growth habit of the NaCl crystal revealed by microscope images (Fig. S4). ResAM treats the crystal to be spherical and positioned in the particle center. However, the dendritic crystals resemble much more an ensemble of narrow cylinders with a surface area much larger than that of a compact sphere. Thus, the diffusion of $Na^+$ and $Cl^-$ ions to the dendritic surfaces is much faster than that to a sphere. The resulting differences can be approximately corrected by choosing an effective diffusion coefficient that compensates the surface ratio.

The number of $Na^+$ and $Cl^-$ ions absorbing onto (or desorbing from) the dendritic cylinders per time unit (i.e. their diffusive flux integrated over the dendritic surface) is

$$\frac{dN_{\mathrm{Na^+/Cl^-}}}{dt} \approx -2\pi\rho_c l_c \times c_{\mathrm{Na^+/Cl^-}} \times D_{\ell,\mathrm{Na^+/Cl^-}} \frac{\partial}{\partial\rho} \ln a_{\mathrm{Na^+/Cl^-}}\bigg|_{\rho\to\rho_c} , \tag{S12}$$

where $\rho$ is the radial distance in cylindrical coordinates, $\rho_c$ and $l_c$ are the mean radius and total length of the dendritic cylinders. For comparison, the diffusive flux of the ions to a centered sphere is

$$\frac{dN_{\mathrm{Na^+/Cl^-}}}{dt} = -4\pi r_s^2 \times c_{\mathrm{Na^+/Cl^-}} \times D^*_{\ell,\mathrm{Na^+/Cl^-}} \frac{\partial}{\partial r} \ln a_{\mathrm{Na^+/Cl^-}}\bigg|_{r\to r_s} , \tag{S13}$$

where $r_s$ is the radius of the compact sphere with the same volume as the dendrites. We introduce effective diffusion coefficients of $Na^+$ and $Cl^-$ ions (marked by a star), $D^*_{\ell,\mathrm{Na^+}}$ and $D^*_{\ell,\mathrm{Cl^-}}$, which compensate the different shapes. Assuming the same activity difference between solid and liquid for both shapes, we obtain:

$$\rho_c \frac{\partial}{\partial\rho} \ln a_{\mathrm{Na^+/Cl^-}}\bigg|_{\rho\to\rho_c} \approx r_s \frac{\partial}{\partial r} \ln a_{\mathrm{Na^+/Cl^-}}\bigg|_{r\to r_s} . \tag{S14}$$

Equating Eqs. S12 and S13 and considering Eq. S14 yields

$$D^*_\ell \approx D_\ell \frac{l_c}{2r_s} . \tag{S15}$$

From the microscope images (Fig. S4), the total length of the dendrites in the droplet with radius of 20 µm is 340 µm. Assuming the thickness of the dendrites is independent of the aerosol particles size, the length of the dendrites is then proportional to the volume of the aerosol particle. The length of the dendrites with radius $R_{\mathrm{ptcl}}$ can be estimated as

$$l_c \approx \max\left(\frac{340}{20^3}\ \mu\mathrm{m}^{-2}\ R^3_{\mathrm{ptcl}}\ ,\ 2 \times r_s\right) , \tag{S16}$$

where the second term in the maximum expression $(2 \times r_s)$ becomes important for small particles and ensures that $D^*_\ell \geq D_\ell$. Typical enhancement factors $D^*_\ell / D_\ell$ vary from 1 for $R_{\mathrm{ptcl}} = 1$ µm to 4 for 10 µm to 400 for 100 µm.

We next derive the flux of ions across the solid/liquid interface, which constitutes the boundary condition for the Nernst-Planck equation. In this case, the electric force can be ignored, since the

$Na^+$ ions and $Cl^-$ ions must always diffuse to the crystal with the same rate, provoking no charge separation. Assuming the uniform activity coefficients in the shell adjacent to the solid crystal, an analytical solution for $j_{Na^+/Cl^-}$ can be obtained:

$$j_{Na^+/Cl^-} = -4\pi D^*_{\ell,Na^+/Cl^-} \times r_s \frac{r_1 + r_s}{r_1 - r_s} \times \Big( c_{Na^+/Cl^-}(r_1) - c_{Na^+/Cl^-}(r_s) \Big) \quad . \tag{S17}$$

Here, $r_1$ is the outer radius of the innermost liquid shell. The $Na^+$ and $Cl^-$ concentrations at $r_1$ are determined from the Nernst-Planck equation of the liquid phase diffusion. At the crystal surface $r_s$, the solution is in equilibrium with the crystal, i.e. $a_{NaCl,solid} = a_{Na^+} a_{Cl^-}$. The activity product of solid NaCl, $a_{NaCl}$, is given in Table S2. Setting $j_{Na^+} = j_{Cl^-}$ and considering the solid equilibrium at the solid surface, the concentrations $c_{Na^+}$, $c_{Cl^-}$ at $r_s$ can be calculated, thus also the flux of $Na^+$ and $Cl^-$ ions from/to the crystal.

##### *Summary of diffusion coefficients in expiratory particles*

The liquid phase diffusion coefficients at infinite dilution in water are summarized in Table S3. The liquid phase diffusion coefficients of $Na^+$ and $Cl^-$ ions, $D_{\ell,Na^+}$ and $D_{\ell,Cl^-}$ at RH 50 - 70% are derived using the EDB data (Fig. S7). The very rapid mass loss immediately after nucleation of the NaCl crystal indicates rapid diffusion of $H_2O$ (Fig. S7B,D number ④ ), while the further crystal growth due to diffusion of $Na^+$ and $Cl^-$ ions is slow (Fig. S7B-D numbers ② and ③ ).

We assume that the liquid-phase diffusion coefficients of all neutral species ($NH_3$, $CO_2$, $CH_3COOH$, $NH_4CH_3COO$, etc.) have the values provided in Table S3 for infinitely diluted solutions and follow the same water activity dependence as $H_2O$ molecules in more concentrated solutions ($D_{\ell,H_2O}(a_w)$). Similarly, the diffusion coefficients of all ions (${NH_4}^+$, ${NO_3}^-$, $CH_3COO^-$, ${HCO_3}^-$, etc.) have the values provided in Table S3 for infinitely diluted solutions and follow the same water activity dependence of $Na^+$ and $Cl^-$ ions in more concentrated solutions, as shown in Fig. S7.

In addition, we assumed that all diffusion coefficients have the same temperature dependence as that of sucrose (55). The error introduced by this approximation is small, because the temperature range relevant for exhalation under indoor conditions is narrow (293.15 K to 307.15 K). The particle temperature of the exhaled particles is calculated considering the latent heat of the evaporating water.

In summary, the use of the Nernst-Planck equation ensures a physically consistent treatment of ion diffusion inside the expiratory particles. The $H^+$ ions participate in establishing charge neutrality, which affects the pH. In ResAM we treat the diffusion problem using the Nernst-Planck equation with $D_{\ell,Cl^-} \approx 1.5 \times D_{\ell,Na^+}$ and adjust all anion and cation diffusivities according to Table S3. We compared these runs with runs assuming $D_{\ell,Na^+} = D_{\ell,Cl^-}$ (with concentrations gradients instead activity gradients) and find pH differences of up to 1 pH unit during times of highest pH gradients (around 1 s in the expiratory plume, see Fig. S21). This demonstrates the importance of different anion-cation diffusivities and their treatment in the Nernst-Planck formulation, which cannot be ignored in the present application.

#### Further ResAM details III: simulation of virus inactivation

##### *Mixing of the exhaled aerosol with indoor air*

In our analysis, we do not consider spatial heterogeneities in the gas-phase concentration of water vapor and gas-phase precursors, but rather a mean field of these parameters and their gas-to-particle transfer controlled by diffusion; this is equivalent to focusing on the virus-containing aerosol outside of the "puff phase" (order of a few seconds), where acidification of the particles would have not occurred to a substantial degree (72–74). The particles considered are persistent and can remain airborne for many minutes to hours, given that the relevant particles have diameters of 5 µm and less (72), hence small settling velocities and gas-to-particle mass transfer in the vicinity of particles controlled by molecular diffusion (Stokes Regime). The gas phase compositions of exhaled air, typical indoor air and various indoor scenarios are provided in Table S4. In our model simulations we use 20°C (293.15 K) as typical indoor temperature and 34°C (307.15 K) as initial temperature of exhaled air (75). We simplified the exhaled air plume as a two-dimensional air flow. The dispersion of the exhaled plume is then approximated using the mean square diffusion length in a 2-D system(76):

$$< r^2 > = 4 \ D_{g,\mathrm{eddy}}\, t. \quad \text{[S18]}$$

$D_{g,\mathrm{eddy}}$ is the turbulent eddy diffusion coefficient of the air inside the room under consideration, which we calculated following Shao and coworkers. They showed that $D_{g,\mathrm{eddy}}$ is a function of the air exchange per hour (ACH) used during indoor mixing ventilation, i.e. $D_{g,\mathrm{eddy}} = f(\mathrm{ACH})$(33). The cross section of the exhaled air plume with an initial area $A(0)$ is then given by

$$A(t) = A(0) + \pi < r^2 >= A(0) + 4\pi D_{g,\mathrm{eddy}} t, \quad \text{[S19]}$$

and the corresponding gas phase mole fraction or mixing ratio ($\chi(t)$) in the diluting exhaled plume including $H_2O$ is

$$\chi(t) = \frac{A(0)}{A(t)} \chi_{\text{exhaled air}} + \frac{A(t) - A(0)}{A(t)} \chi_{\text{indoor air}}. \quad \text{[S20]}$$

The relative humidity is then the ratio of partial pressure (i.e., the total pressures × $H_2O$ mole fraction) over the vapor pressure of pure water(77). For normal breathing, we assume an initial area $A(0) = 2 \times \pi \times 0.75^2$ cm$^2$ (corresponding to the size of two nostrils with radius of 0.75 cm). Shao et al. (33) report $D_{g,\mathrm{eddy}}$ values for an ACH range from 0.43 to 2.97 based on their experiments using toluene and acetone. The temperature of the exhaled air plume is calculated analogously to the gas phase mixing ratio (see Eq. S20).

For ACH = 2, we use $D_{g,\mathrm{eddy}} = 50$ cm$^2$/s in the present simulations. For ACH = 0.1, we use the lower limit $D_{g,\mathrm{eddy}} = 10$ cm$^2$/s (33). We note that $D_{g,\mathrm{eddy}}$ is roughly proportional to ACH$^2$ (see Table 3 of Shao et al.(33)). Thus, we extrapolate $D_{g,\mathrm{eddy}}$, and obtain $D_{g,\mathrm{eddy}} = 1250$ cm$^2$/s for ACH = 10.

The exhaled aerosol exchanges semi-volatile species with the gas phase. Nazaroff and Weschler reviewed the indoor air acids and bases (25). The gas phase composition is given by Table S4. By adding $HNO_3$ to the indoor air, the partitioning of $NH_3$ will change: the lower pH of background indoor aerosol allows more uptake of $NH_3$, i.e. $NH_3$ is scavenged by the background aerosol (see again Table S4). The reduction of $NH_3$ by adding $HNO_3$ to the indoor is calculated assuming an aerosol mass concentration of 20 µg/m$^3$, which is a typical value for household indoor air (78). We use a simplified composition of background aerosol of aqueous sucrose/sulfate/$Cl^-$/$NO_3^-$ solution. Our final results are not strongly dependent on aerosol mass concentration or composition. Under typical indoor conditions, 20°C and 50% RH, the gas phase $NH_3$ is reduced from 36.3 ppb to 1.88 and 0.376 ppb, by adding 10 ppb and 50 ppb $HNO_3$ to the indoor air, respectively.

The composition and pH of the exhaled aerosol is calculated for sizes ranging from 0.2 µm to 1000 µm. In combination with the measured inactivation time of virus at given pH, the 99%-inactivation time of viruses in the exhaled aerosol can then be readily calculated. The resulting composition, pH and virus concentration in exhaled aerosol are shown in Fig. 4 for 50% RH and by Fig. S8 by for 80% RH.

##### *Virus inactivation times and residual infectious fractions in aerosol particles*

In the aerosol phase, the extent of virus inactivation in a microscopic particle depends on particle size, pH-dependent inactivation processes, and the time since exhalation. As prerequisite for the treatment of the microscopic aerosol problem, we use our macroscopic inactivation measurements shown in Fig. 1 (based on matrix volumes between 10 µl and 1 ml), to derive 1/e-inactivation times, $\tau$, for all three viruses. These bulk measurements provide $\tau$ as a function of pH and of the mass fraction of solutes, $x_m$, which we fit for convenience to arctan functions. For IAV, we approximate the inactivation times by the following fit:

$$\begin{aligned}\tau(\mathrm{pH}, x_m)/\mathrm{s} = &\exp(-2.045 + 0.46916 \times \mathrm{pH}) + \\ &\exp\Big[5.0131 + 6.2214 \times \arctan\Big(1.2972 \times \\ &(\mathrm{pH} - 5.41836 + 1.4134 \times x_m) \times (1.012728 - 0.4714 \times x_m)\Big)\Big]. \end{aligned} \quad \text{[S21]}$$

Based on Eq. S21, Fig. 1 shows the calculated 99%-inactivation times for different solute mass fractions (blue curves).

The inactivation time of SARS-CoV-2, which shows no strong dependence on $x_m$ over the measured range (see Fig. 1), is expressed as a function of pH by

$$\tau(\mathrm{pH})/\mathrm{s} = \exp\Big[4.25622 + 5 \arctan\Big(2.0308 \times (\mathrm{pH} - 2.1729)\Big)\Big]. \quad \text{[S22]}$$

Finally, the inactivation time of HCoV-229E as a function of pH is expressed by

$$\tau(\mathrm{pH})/\mathrm{s} = \exp\Big[8.3045 + 3.3891 \arctan\Big(0.50961 \times (\mathrm{pH} - 2.2457)\Big)\Big]. \quad \text{[S23]}$$

Next, we determine $N(R_0, t)/N(R_0, 0)$, which is the residual infectious virus fraction at time $t$ in an aerosol particle exhaled at time $t = 0$ with initial radius $R_0$ . In shell $k$, the infectious virus fraction is given by

$$\ln \frac{N_k(R_0, t)}{N_k(R_0, 0)} = -\int_0^t \frac{dt'}{\tau(\mathrm{pH}_k(t'), x_{m,k}(t'))} \; . \quad \text{[S24]}$$

Here, $N_k(R_0, 0)$ and $N_k(R_0, t)$ are the number of infectious viruses in shell $k$ at time 0 and time $t$, respectively, and $\mathrm{pH}_k(t')$ and $x_{m,k}(t')$ are the simulated pH and mass fraction of solutes of shell $k$ at time $t'$. For the entire particles, the infectious fraction of virus, $N(R_0, t)/N(R_0, 0)$, is given by

$$\frac{N(R_0, t)}{N(R_0, 0)} = \frac{\sum_k N_k(R_0, t)}{\sum_k N_k(R_0, 0)}. \quad \text{[S25]}$$

Note that the summation over shells in Eq. S25 results in a volume-weighted averaging for the inactivation times. The times for e-folding, $10^{-2}$- and $10^{-4}$-fold reductions calculated from Eq. S25 as a function of initial particle radius $R_0$ are shown in Fig. 4.

#### Modeling of indoor airborne viral load and relative risks of transmission

The airborne load of infectious viruses depends on the emission rate of these viruses on the one hand (source), and the deposition of aerosol particles, the ventilation of indoor air, and finally the inactivation of viruses in aerosol particles on the other hand (sinks). Assuming well-mixed ventilation, the airborne viral load (number of infectious viruses per volume of air) can be expressed as

$$\bar{n}_{\text{virus}}(t) = \frac{1}{V}\int_0^t \int_{50\,\text{nm}}^{\infty} \nu(R_0)\,\frac{dQ}{d\log(R_0)}\,f_{\text{depo}}(t-t')\,e^{-\text{ACH}\times(t-t')}\,\frac{N(R_0,t-t')}{N(R_0,0)}\,d\log(R_0)dt'. \quad \text{[S26]}$$

Here , $V$ is the room volume occupied per infected person, $dQ/d\log(R_0)$ is the aerosol number emission rate distribution of one human (see Fig. 4E), $\nu(R_0)$ is the number of virus in one aerosol particle of radius $R_0$ and $N(R_0,t-t')/N(R_0,0)$ is the residual infectious virus fraction (see Eq. S25). (In passing we note that $dQ/d\log(R)$ in Fig. 4E refers to $t=1$ s, which we take into account by extrapolating backward to $t=0$ in Eq. S26.) Finally, $f_{\text{depo}}(t-t')$ is the reduction factor due to deposition, given by

$$f_{\text{depo}}(t-t') = e^{-\int_0^{t-t'} \frac{v_{\text{dep}}(R(t''))}{h} dt''}, \quad \text{[S27]}$$

where $v_{\text{dep}}(R)$ is the deposition velocity of aerosol particles with radus $R$, and $h$ is the height of the room.

We assume $V$ is 10 $m^3$ per infected individual, and the height of room $h$ is 2 meters, which represents a typical length scale for deposition (assuming the indoor air to be well-mixed due to convection and turbulence and irrespective of the particular position of the infected human). The emission rates of exhaled aerosol during breathing, coughing and speaking/singing shown in Fig. 4E from Pöhlker et al. (36) are used. We calculate the particle deposition velocity $v_{\text{dep}}$ from experimentally determined dry deposition data (Seinfeld and Pandis, 2006; Fig. 19.2, data for $u^* = 44$ cm/s(79)). A slower deposition velocity ($u^* = 11$ cm/s) results in an even stronger decrease in the relative risks of transmission. The steady state airborne viral load can be calculated by integrating Eq. S26 over a sufficiently long time $t$, such that the viral load converges to a constant value, $\bar{n}_{\text{virus}}(t \to \infty)$ .

Results for $\nu(r) \equiv 1$, i.e. each aerosol particle containing one virus, are shown in Figs. 5 and S15. Results for $\nu(r) \propto r$, i.e. virus concentration that are proportional to the size of the aerosol particles, are shown in Fig. S16. Also for $\nu(r) \propto r$, significant reductions of transmission risks can be achieved by air acidification for all kinds of emissions (breath, cough, speak/sing).

#### ResAM sensitivity studies

We performed sensitivity analyses for the modeled inactivation times of IAV and SARS-CoV-2 by systematically varying eight critical model parameters (Figs. S11 and S12. The following listing by capital letters refers to the sensitivity model runs depicted in the various panels of these figures:

**A: Sensitivity to initial pH buffer strength of exhaled aerosol.** In the model runs presented in this paper, the initial pH during particle formation is 6.6. Sensitivity tests were performed with ResAM for initial pH of 6.2 or 7.0 (see subsection "Aerosol initial pH buffering" for details). An initial pH of 7.0 leads to slower, and an initial pH of 6.2 to faster virus inactivation for particles with radii > 1 µm, in particular in acidified air. The effect of initial pH is less important for smaller particles, which are dominant during exhalation. Note that the sensitivity test for pH 7.0 covers also the case with original SLF concentrations initially in equilibrium with 46 500 ppm $CO_2$.

**B: Sensitivity to mixing speed of exhaled air plume with indoor air.** A longer mixing time, i.e. a smaller eddy diffusion coefficient $D_{g,\mathrm{eddy}}$ (see Eq. S18), has only a weak effect, leading to slightly longer inactivation times.

**C: Sensitivity to uncertainties in liquid phase diffusion coefficients.** The exact value of the liquid phase diffusion coefficient plays a critical role for aerosol particles with radius larger than about 1 µm. The impact becomes negligible for small particles, when inactivation times are short anyway and are governed by the mixing with the indoor air (via $D_{g,\mathrm{eddy}}$) or by the virus inactivation time.

**D: Sensitivity to $NH_3$ gas phase mixing ratio.** In typical indoor air, an increase in the gas phase $NH_3$ mixing ratio increases the inactivation time negligibly. Conversely, a nearly complete removal of $NH_3$ decreases the inactivation time only for exhaled particles with radii > 1 µm, but also this becomes negligible when the air is enriched with $HNO_3$.

**E: Sensitivity to $HNO_3$ gas phase mixing ratio.** In typical indoor air, the inactivation time of IAV changes only slightly, when the $HNO_3$ gas phase concentration varies by a factor of 2, and only for exhaled aerosol particles with radius > 1 µm.

**F: Sensitivity to indoor temperature.** Inactivation times slightly decrease with increasing temperature. Neither the temperature dependence of the different physicochemical model parameters, nor that of virus inactivation kinetics, critically affect the inactivation times.

**G: Sensitivity to relative humidity.** The higher the relative humidity, the shorter the inactivation time. However, the differences remain small in the RH range 40 - 80%.

**H: Sensitivity to aerosol matrix composition.** Depending on the inorganic/organic ratio, the activity coefficients may differ, which in turn affects virus inactivation times. Assuming unchanged liquid phase diffusion coefficients, we performed sensitivity runs reducing the composition to only the organic components of SLF or to pure NaCl. The computed inactivation times typically differ by less than 50% from those of SLF.

#### Comparison of published inactivation data with ResAM simulation

Figures S13 and S14 present a comparison of the 99%-inactivation times computed by ResAM (highlighted in green) with published measurements (colored circles). Previous work on IAV and SARS-CoV-2 in aerosol particles has focused on the dependence on RH, but did not consider the role of other air constituents that may modulate aerosol pH. The published data for IAV suggest a weak decrease of 99%-inactivation times with RH (Fig. S13). This trend is also captured by ResAM, and can be attributed to the increasing liquid phase diffusivity at higher RH. ResAM computation furthermore reveals a strong dependence of IAV inactivation time on air composition, and to a lesser extent on particle radius. Air filtration, e.g., by HEPA filters, can be expected to partly remove both organic and inorganic acids and bases, leading to a net increase in aerosol pH, and hence to prolonged inactivation times. In contrast, air conditioning by $HNO_3$ leads to aerosol acidification and shorter inactivation times. Published inactivation data for aerosolized IAV are consistent with ResAM estimates, assuming that experiments were conducted in partially purified air. Air treatment prior to experimentation in rotating drums was described in two studies (8, 9), and includes HEPA filtration and hydrocarbon traps. While earlier studies did not specify if air treatment took place, we can assume that all rotating drum experiments use filtered air to minimize contamination by ambient aerosol particles. According to ResAM model predictions, these treatments typically resulted in air purification of 80-99%.

SARS-CoV-2 exhibits little dependence on RH, as seen in both published data and in ResAM computations (Fig. S14). For this virus, ResAM determines only small variations in inactivation times due to air purification, because SARS-CoV-2 is insensitive to the ensuing changes in aerosol pH (Fig. 4). When air is amended with $HNO_3$, however, the resulting increase in aerosol acidity causes inactivation times shift to lower values. A comparison of inactivation times between published data and ResAM estimates is inconclusive. Due to the high stability of SARS-CoV-2 in unconditioned air, inactivation kinetics are difficult to measure in short-term rotating drum experiments. Consequently, inactivation rate constants exhibit large variability and inactivation times span over a wide range, even when determined under identical experimental conditions. Nevertheless, we observe that published inactivation times coincide with ResAM model results for typical room air all the way to fully purified air.

We conclude that ResAM is capable of reproducing inactivation trends of IAV and SARS-CoV-2 as a function of RH, as well as inactivation times under reasonable assumptions of the experimental conditions used. We furthermore note that in particular for pH-sensitive viruses, knowledge of air composition is critical to interpret and reproduce experimental results.

#### Investigation of $CH_3COOH$ as potential agent against airborne viruses

##### *Acetic acid enrichment does not enhance virus inactivation times*

Based on an ancient medical custom (80), the boiling of vinegar as preventive measure against airborne pathogens has a long tradition, even though it remains unclear whether there is a scientific basis or whether it is folk tale. In February 2003, during the early phase of the Severe acute respiratory syndrome (SARS) outbreak in China, local media listed some preventive measures, including enhanced ventilation or using vinegar fumes to "disinfect the air", leading to vinegar being sold out in some cities (22). Diluted household vinegar containing 0.4-0.8 wt% acetic acid ($CH_3COOH$) was shown to reduce the infectivity of Human Influenza A/H1N1 in tissue culture by 7 orders of magnitude in less than 1 minute (81), and even against SARS-CoV-2 there is evidence of ($CH_3COOH$) being antiviral (82). However, the results of these in vitro experiments, which use well-mixed bulk volumes with a prescribed pH, are not directly applicable to aerosol particles freely floating in a gas. Whether a substance, such as $CH_3COOH$, readily partitions from the gas to the particulate phase depends on its volatility and on whether it may lead to sufficiently fast virus inactivation inside the particles. This, in turn, depends on a variety of aerosol properties and processes, which determine the disinfecting power of the substance, e.g., by attaining a high acidity or oxidation potential. Indeed, our measurements of $CH_3COOH$ vapor pressure (see below) and model simulations (see Fig. S9) demonstrate that $CH_3COOH$ is too volatile and a too weak acid to enhance virus inactivation, at least at concentrations below the permissible exposure level of 10 ppm.

##### ***Measurements of vapor pressure of ammonium acetate***

To estimate the partitioning of gas phase acetic acid and ammonia in equilibrium with condensed phase ammonium acetate we measured evaporation rates of single, aqueous ammonium acetate particles levitated in the EDB. Details of the technique and data analysis were previously described (e.g. (83, 84)). Briefly, an aqueous ammonium acetate particle (ca. 15 µm radius) was injected into the EDB at a temperature of 20.4°C and 87.5% RH, and its mass evaporation rate was measured by the voltage compensating the particle's gravitational force. Analyzing nineteen such experiments yielded a vapor pressure of 0.12±0.05 Pa for ammonia and of 0.062±0.010 Pa for acetic acid.

### Supplementary Figures

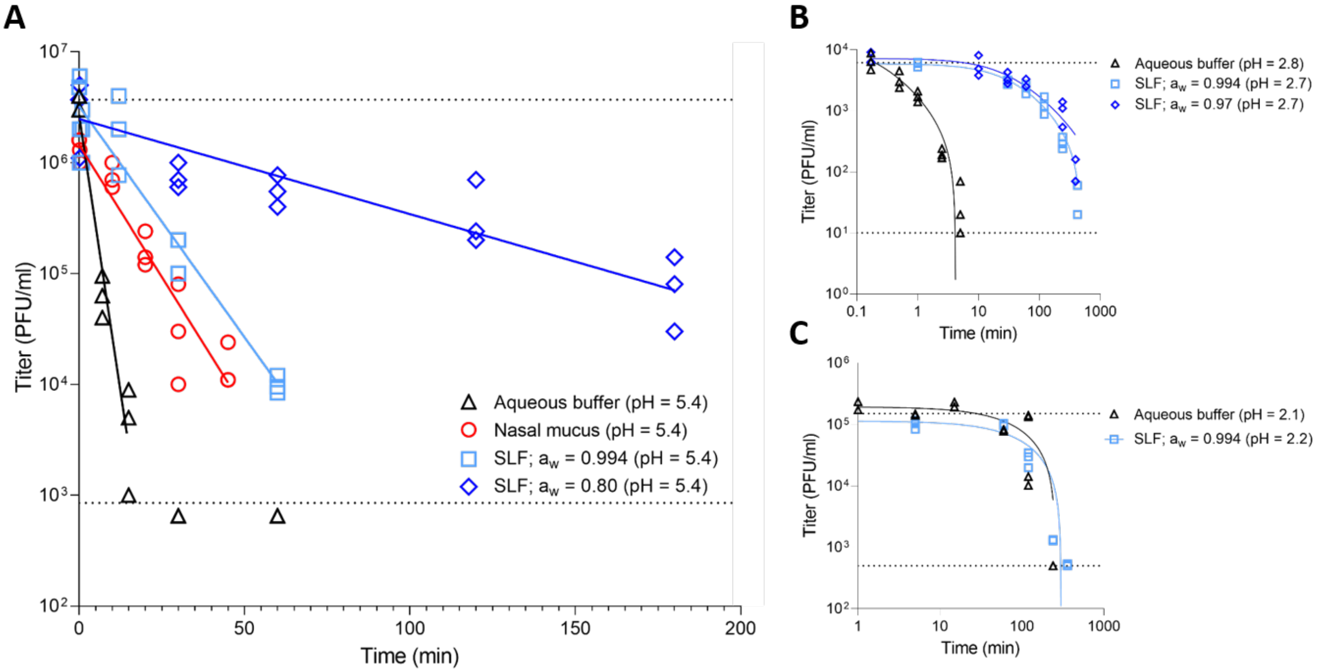


**Fig. S1. Examplary inactivation curves of IAV, HCoV-229E and SARS-CoV-2 measured in aqueous buffer, in nasal mucus, and in different synthetic lung fluid (SLF) concentrations.** (**A**) IAV, (**B**) SARS-CoV-2, and (**C**) HCoV-229E titers as a function of exposure time at a pH of 5.4, 2.7 - 2.8, or 2.1 - 2.2, respectively. Individual data points indicate triplicate measurements at each time-point. Solid lines represent model fits assuming first-order decay. Dashed lines correspond to the average starting titer (top line) and detection limit (bottom line) for each respective virus.

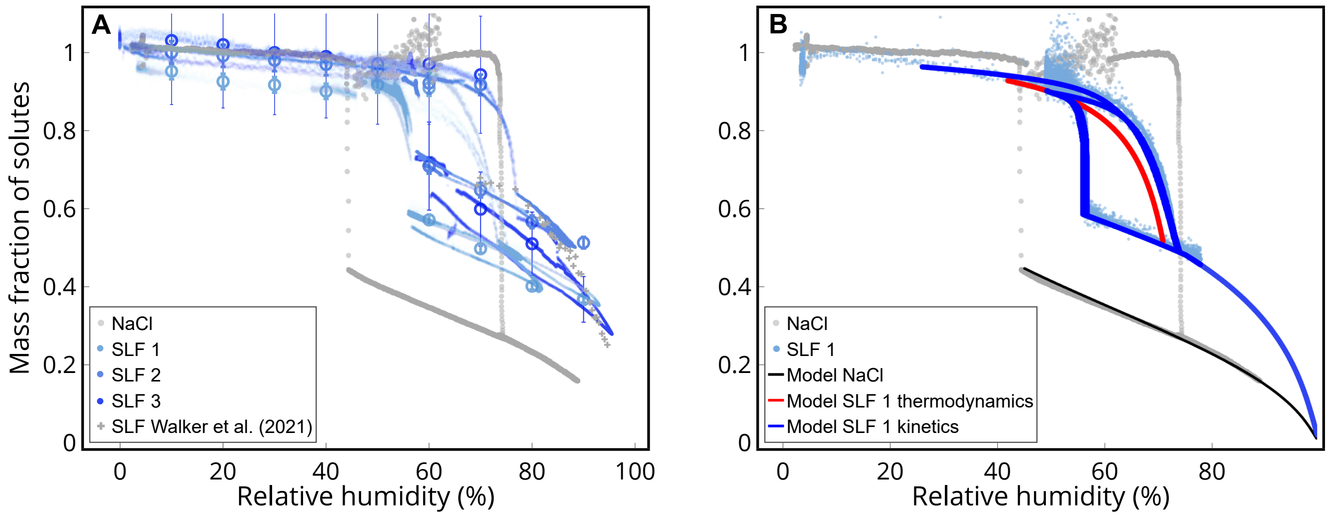


**Fig. S2. Thermodynamic equilibrium properties of synthetic lung fluid (SLF).** (**A**) Voltage data obtained from EDB measurements, converted to mass fraction of solutes (MFS), shown for one NaCl particle (NaCl - grey dots) and three different SLF particles (SLF 1 - light blue dots, shown also in Fig. 2, SLF 2 - medium blue dots, and SLF 3 - dark blue dots) during deliquescence/efflorescence cycles. Light grey crosses indicate measurements of SLF particles by Walker et al. (85). Open circles indicate the average for wet and dry conditions with error bars indicating the systematic error of the measurement (± 0.5 V). The hygroscopicity of the SLF particles are significantly smaller than of aqueous NaCl particles, but show large variability between different particles (beyond measurement uncertainty) owing to the large degree of multi-scale granularity of natural or synthetic lung fluids (see TEM images in Fig. S5). Note that MFS above 1 are an artifact due to very small changes in voltage under very dry conditions resulting in very large changes in MFS. (**B**) The thermodynamic equilibrium properties of the biophysical model ResAM were constrained using the data in **A**. Grey dots indicate NaCl as in **A**. Light blue dots indicate SLF 1 (as in **A** but only cycles 2 and 3). Curves show model results of ResAM. The black curve indicates modeled pure NaCl solution. The lower branch of the red solid curve indicates thermodynamic equilibrium behavior of SLF as function of RH, calculated by adjusting the activity of the organic fraction of SLF, $a_{\mathrm{org}} = f(a_w) = f(\mathrm{RH})$ to fit the EDB measurements. The upper branch of the red solid curve is the deliquescence curve of SLF in thermodynamic equilibrium as calculated by ResAM, which fits the experimental data reasonably well, but fails to accurately describe the steep portion of the deliquescence branch. The dark blue curves indicate a fully kinetic simulation with ResAM, which shows that only taking the slow diffusive processes into account provides full overlap with measurements during the two hours long deliquescence process.

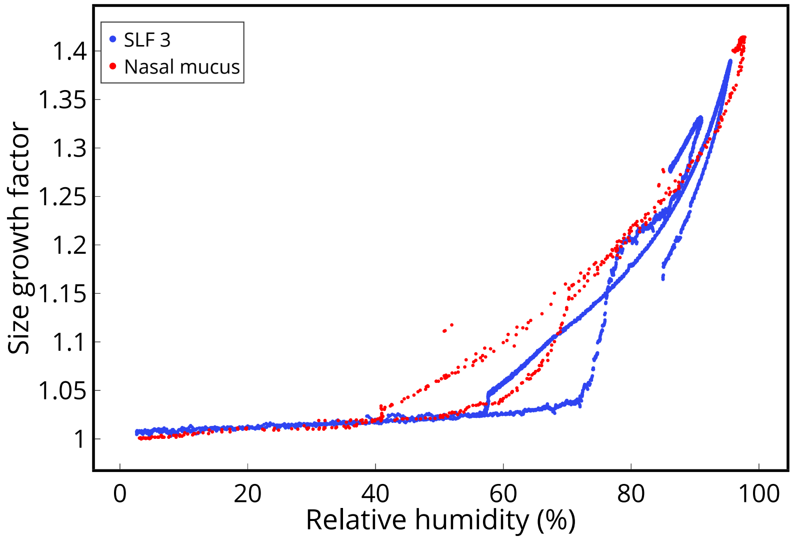


**Fig. S3. Size growth factor of SLF and nasal mucus.** Mie-Resonance spectra (83, 84) of two selected EDB measurements of SLF (blue data points, particle SLF 3 in Fig. S2) and nasal mucus (red data points) have been normalized to show the size growth factor as function of RH. Size growth at RH $>$ 80% is similar for both particles, but efflorescence and deliquescence RHs differ. For SLF particles, salt effloresces at $\sim$ 56% RH, whilst the nasal mucus particle effloresces only at 42% RH. In addition, deliquescence occurs in SLF around 75% RH, similar to aqueous NaCl, whereas nasal mucus deliquesces at RH $\sim$ 69%. This indicates that SLF particles are dominated by NaCl, whereas the nasal mucus contains significant amounts of other salts.

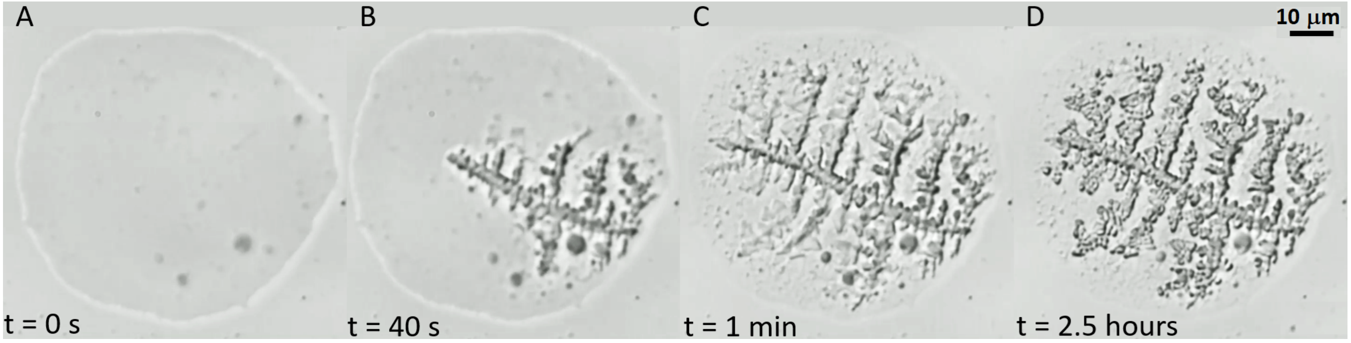


**Fig. S4. Time series of microscope images (50x) of an efflorescing synthetic lung fluid (SLF) particle deposited on a hydrophobically coated substrate in a temperature and humidity controlled cell.** The setup is described in detail by Ciobanu et al. (86). At 20.5°C and 54% relative humidity the particle's radius is approximately 40 µm (scalebar represents 10 µm). (**A**) Liquid particle at $t = 0$, when efflorescence begins. (**B**) 40 s after the start of crystallization with rapid dendritic NaCl crystal growth. (**C**) 1 min after initial crystallization the crystal reaches the edge of the droplet, slowing the speed of further crystallization dramatically. (**D**) 2.5 h after initial crystallization the crystal is still growing slowly, leading to a coarsening of the crystal structure. The size of the dendrites is used to calculate the morphology-dependent effective diffusion constant of $Na^+$ and $Cl^-$ ions in the remaining, mainly organic solution (see subsection on liquid-solid interface and dendritic crystal growth habit in the supplementary text).

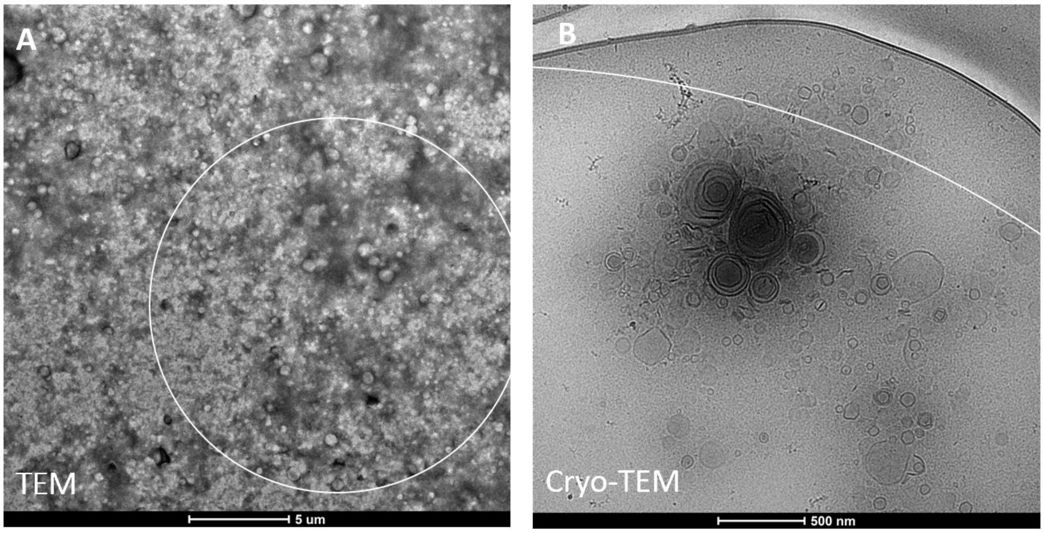


**Fig. S5. Microstructures in synthetic lung fluid (SLF).** The structure of SLF was investigated using (**A**) TEM and (**B**) cryo-TEM. SLF exhibits a colloidal structure, possessing vesicles similar in nature to those found in lung fluid extracts (87). DLS measurements of the particles in SLF showed that the particles have an average particle size of 124 nm, but ranged from 59 nm to 220 nm. The size of liposomes influences the size of particles in the SLF solution. For comparison with the shown inhomogeneities, the white circles indicate typical sizes of particles investigated in the EDB in the present work.

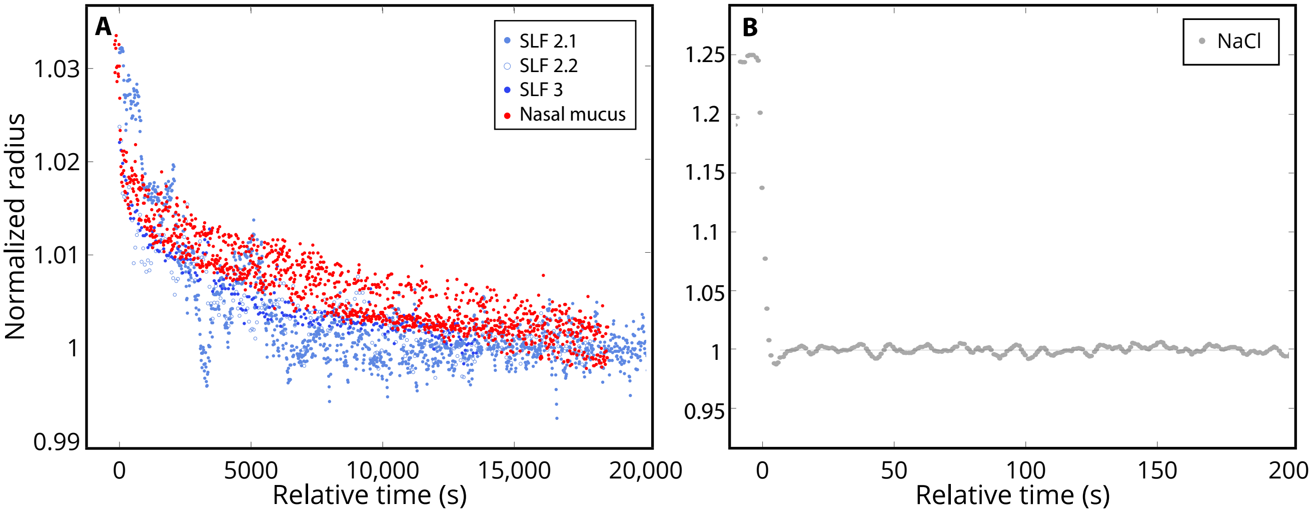


**Fig. S6. Second kinetically impeded stage of efflorescence after the initial rapid first stage of two different SLF particles and one nasal mucus particle.** For comparison, an NaCl particle displays only a single efflorescence stage. (**A**) Similar to Fig. 2B, but focusing on the slow crystal growth stage after the initial fast loss of water. In contrast to Fig. 2B, here the data is obtained from the Mie-Resonance spectra, providing a more accurate (though also much more elaborate) determination of radius than voltage does (83, 84). Light blue symbols show two different efflorescence cycles of particle SLF 2 (labelled as SLF 2.1 and 2.2). Dark blue symbols show measurements for particle SLF 3. Red symbols indicate measurements for a nasal mucus particle. These independent spectroscopic data do not only corroborate the voltage data showing kinetically impeded water loss, but they confirm reproducibility. Most importantly, nasal mucus shows a similar behavior to SLF, i.e. this is not a unique property of SLF. An exponential curve was fitted to these data sets and characteristic times were calculated as: 1500 s (SLF 2.1), 3000 s (SLF 2.2), 1700 s (SLF 3), and 2800 s (nasal mucus). (**B**) Efflorescence of a NaCl particle for comparison (voltage measurements by Braun et al. (88) converted to normalized radius). The NaCl particle loses water almost instantaneously and equilibrates with the environment within a few seconds. Note that the radius cannot be obtained for NaCl particles after efflorescence due to their non-sphericity, therefore voltage data have to be used.

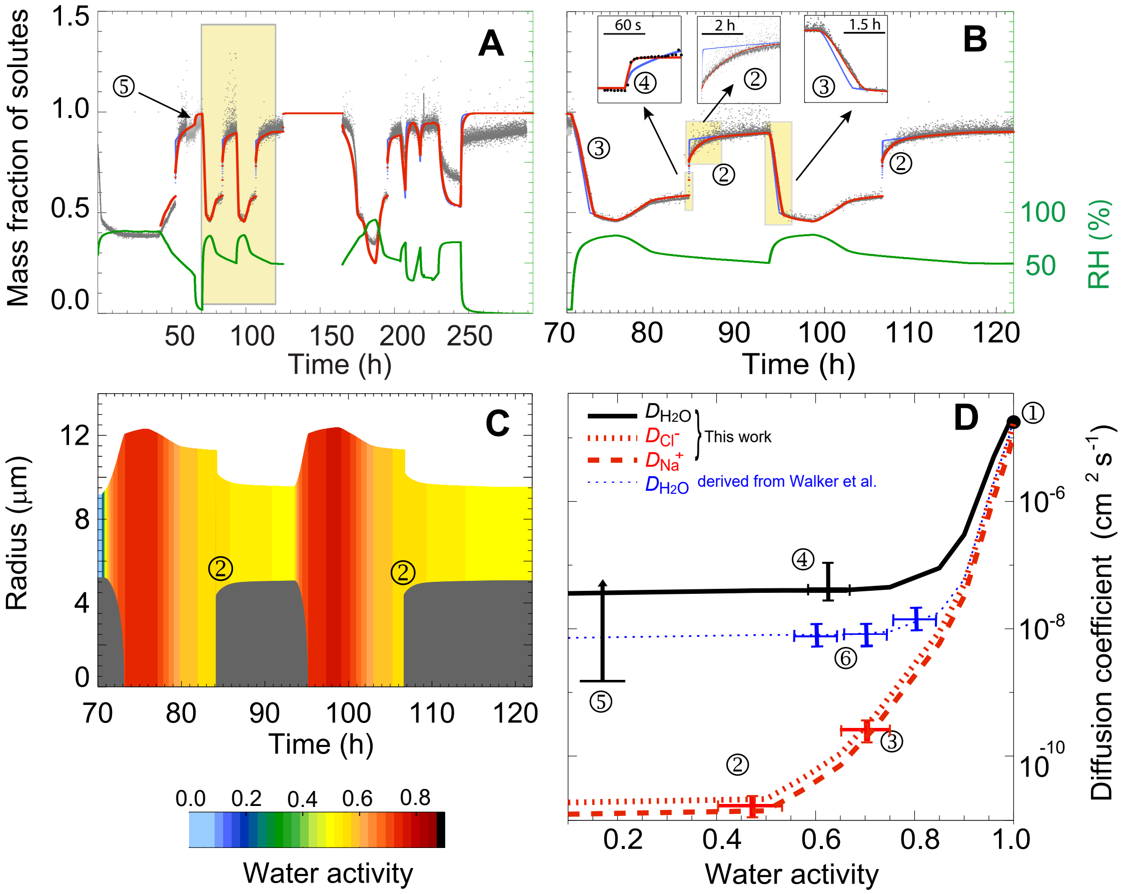


**Fig. S7. Measured and modeled deliquescence/efflorescence cycles of a synthetic lung fluid (SLF) particle levitated in an electrodynamic balance (EDB) and forced by prescribed changes in relative humidity (RH).** (**A**) Similar to Fig. 2, but showing mass fraction of solute (grey dots) instead of voltage, along with prescribed relative humidity, RH (green curve) and ResAM results (red curve). Full 12-day measurement of SLF particle with a dry radius $R$ of $\sim$ 9.7 µm undergoing four efflorescence/deliquescence cycles. Cycle 1 (40-70 h) was used for instrument tuning and camera adjustments. Cycles 2+3 (highlighted in yellow) were used to derive the liquid phase diffusion constants of water molecules and $Na^+$ and $Cl^-$ ions, $D_{\ell,H_2O}$ and $D_{\ell,\mathrm{ions}}$. The period 125-165 h was used to perform independent measurements of the drag force on the particle induced by the gas flow (not shown). Cycle 4 yielded mass fractions of solutes and diffusivities similar to the previous Cycles 2 and 3. (**B**) Zoom on the highlighted Cycles 2+3, showing fast initial crystal growth and loss of water $\sim$ 4 s after efflorescence, followed by slower ($\Delta t \sim$ hours) crystal growth caused by $Na^+$ and $Cl^-$ ions diffusing to the crystal through liquid of progressively increasing viscosity, with a diffusion constant of about $D_{\ell,\mathrm{ions}} \approx R^2/\Delta t \approx 10^{-10}$ cm$^2$/s. Red and blue curves show the results of the respiratory aerosol model ResAM. The red curve indicates the fully fitted model with $D_{\ell,\mathrm{ions}} \ll D_{\ell,H_2O}$ (see black and red curves in panel D for details), whilst the blue curve assumes all diffusivities to be identical ($D_{\ell,\mathrm{ions}} = D_{\ell,H_2O}$) and to follow the blue line in panel D. (**C**) Modeled water activity and NaCl crystal growth (dark grey cores) in the particle during the humidification cycles highlighted in yellow in panel A. (**D**) Diffusion coefficients for $H_2O$, $Na^+$ and $Cl^-$ as derived from the measurements in panels A-C. The black curve indicates $D_{\ell,H_2O}$ in SLF. The red curves indicate $D_{\ell,\mathrm{ions}}$ (with $D_{\ell,Cl^-} \sim 1.4 \times D_{\ell,Na^+}$, see section "Liquid phase diffusion"). The blue curve is an estimate of $D_{\ell,H_2O}$ derived by ResAM from measurements in SLF by Walker et al. (85).

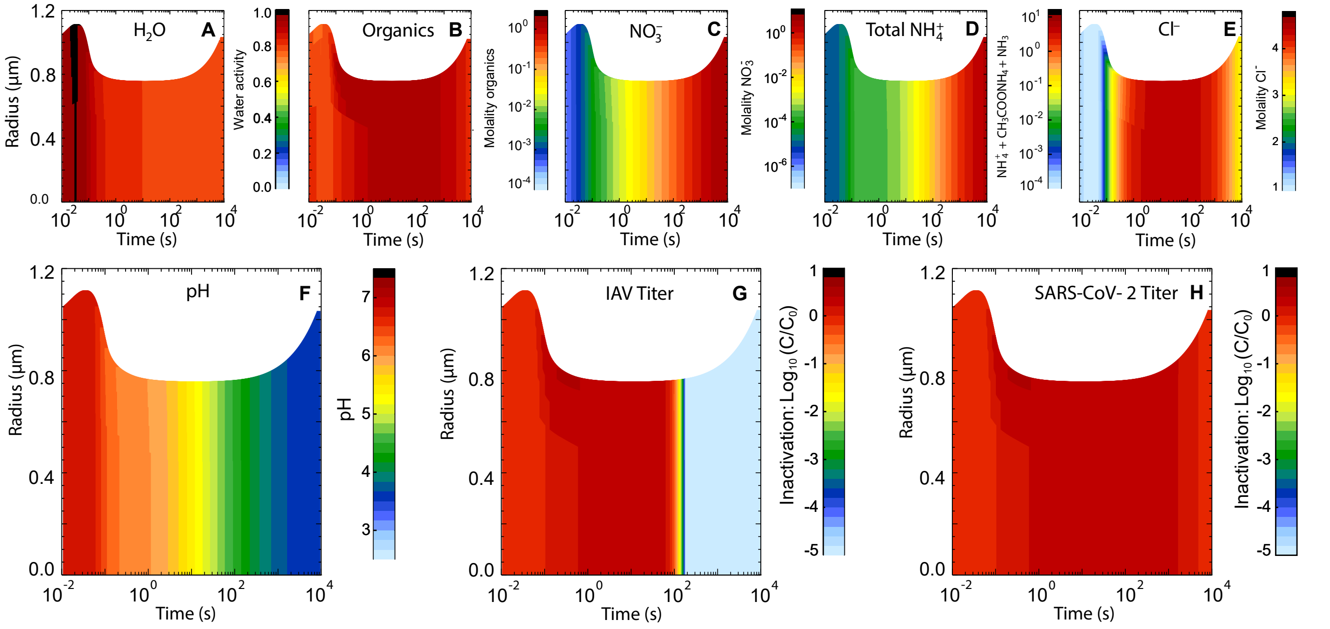


**Fig. S8. Evolution of physicochemical conditions within a respiratory particle and concomitant inactivation of viruses trapped in the particle during transition from nasal to humid indoor air conditions, modeled with ResAM.** Same as Fig. 3 but for 80% RH. As a consequence of the higher RH, the particle shrinks less, concentrations of all chemical species stay lower, and the salt does not effloresce. However, overall the evolution of pH is not very different upon exhalation, leading to an inactivation time of $\sim$ 2.5 minutes for influenza virus at 80% RH compared to $\sim$ 3 minutes at 50% RH. In either case, coronaviruses remain active.

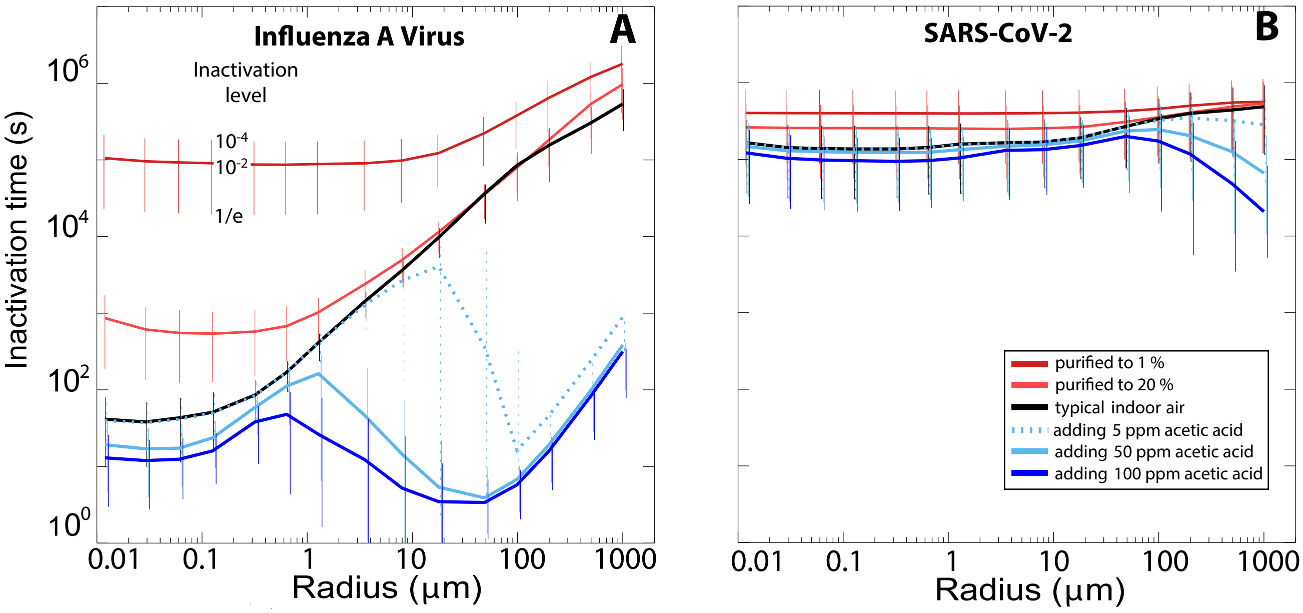


**Fig. S9. Modeled inactivation times for IAV and SARS-CoV-2 as a function of particle size when adding acetic acid to indoor air.** Inactivation times for (**A**) IAV, and (**B**) SARS-CoV-2, when expelled into typical room air (black curve), indoor air purified to 20% (orange curve) or 1% (red curve), or indoor air supplemented with 5 ppm (dotted light blue curve), 50 ppm (solid light blue curve) or 100 ppm (dark blue curve) acetic acid. The exposure threshold of acetic acid is 10 ppm averaged over an 8-hour work shift. Enrichment of acetic acid in indoor air within this acceptable range cannot achieve a sufficient reduction in particle pH and, therefore, yields no robust reduction in virus inactivation times (except for large particles, in which very low pH ($< 2.5$) can be achieved due to much higher gas phase concentration of $CH_3COOH$ than $NH_3$ and the faster liquid diffusivity of undissociated $CH_3COOH$ molecules than $NH_4^+$ ions).

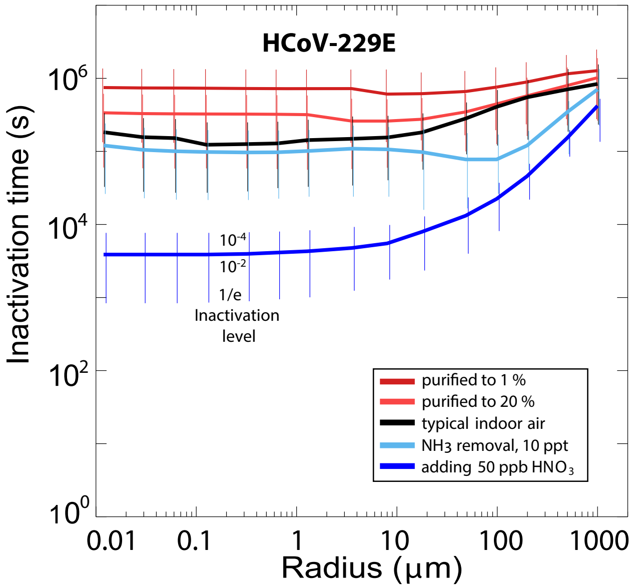


**Fig. S10. Inactivation time for HCoV-229E as function of particle radius and air composition.** Analogous to 4D-E. Indoor air with typical composition (black), enriched with 10 or 50 ppb $HNO_3$ (blue), or purified air with $HNO_3$ and $NH_3$ reduced to 20% or 1% (red). Whiskers show reductions of virus load to $10^{-4}$ (upper end), $10^{-2}$ (intersection with line) and 1/e (lower end). The gas phase compositions of exhaled air and the various cases of indoor air are defined in Table S4. The exhaled air is assumed to mix with the indoor air by a turbulent eddy diffusion coefficient of 50 $cm^2\ s^{-1}$. Radius values refer to the particle size 1 s after exhalation.

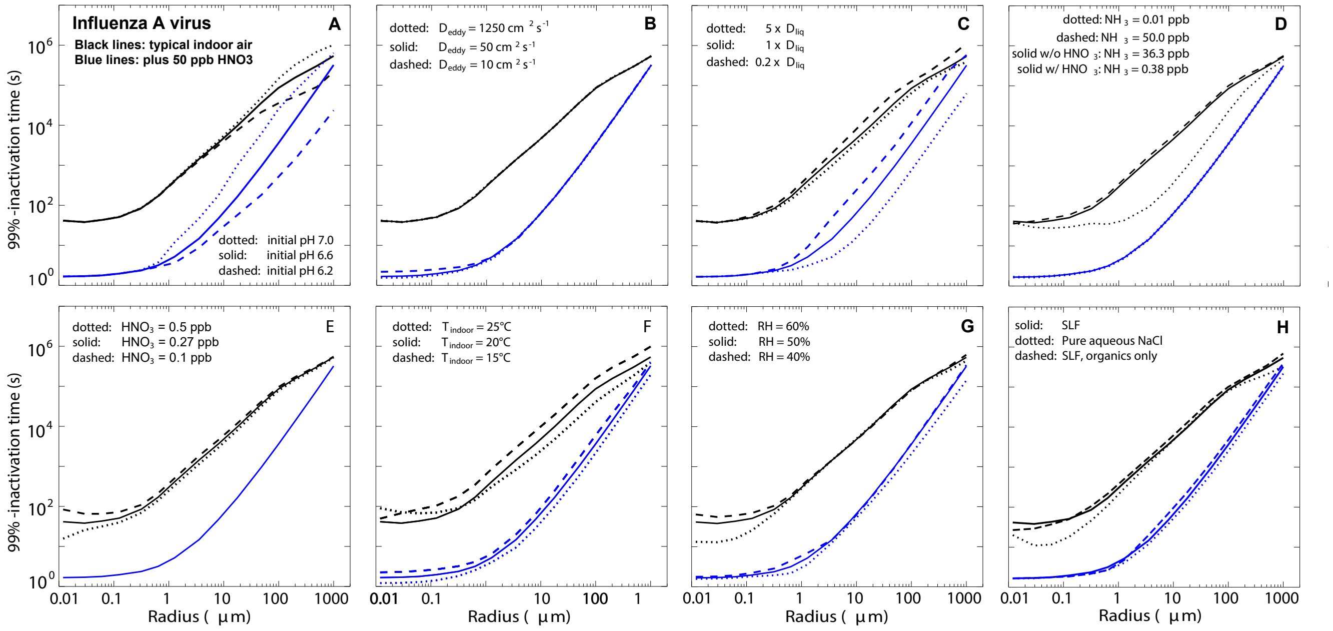


**Fig. S11. Sensitivity studies for the inactivation of IAV.** In each panel, black lines indicate typical (untreated) indoor air, and blue lines indicate typical indoor air supplemented with 50 ppb $HNO_3$. (**A**) Sensitivity to buffering as expressed by initial exhaled particle pH varying from 6.2 to 7.0 (see supplementary text for details). (**B**) Sensitivity to mixing speed of exhalation plume with indoor air, expressed by an eddy diffusion coefficient (see Eq. S20), which is characteristic for a given ventilation (air change per hour, ACH). The highest eddy diffusion coefficient 1250 $cm^2s^{-1}$ applies to ACH = 10, 50 $cm^2s^{-1}$ to ACH = 2 and 10 $cm^2s^{-1}$ to ACH = 0.1. (**C**) Sensitivity to uncertainties in the liquid phase diffusion coefficients (increasing or decreasing by a factor of 5). (**D**) Sensitivity to indoor ammonia concentrations varying from conditions in a clean room (0.01 ppb) to conditions in a polluted house or nursing home (50 ppb)(25). (**E**) Sensitivity to indoor gaseous nitric acid concentrations varying from conditions found in libraries and museums (0.1 ppb) to conditions of the highest indoor values in houses with natural ventilation (0.5 ppb)(25). (**F**) Sensitivity to indoor temperature (15-25°C), applying the temperature dependencies of all physicochemical parameters and assuming the virus inactivation rates to increase by a factor of 1.5 when temperature is increased by 5°C (conservatively estimated based on results in (6)). (**G**) Sensitivity to indoor relative humidity (with 40% RH values obtained from linear extrapolation of all parameters between 60% and 50%, i.e. with high uncertainty). (**H**) Sensitivity to exhaled aerosol composition, namely reducing composition to only the organic components of SLF (dashed line) or pure NaCl (dotted line).

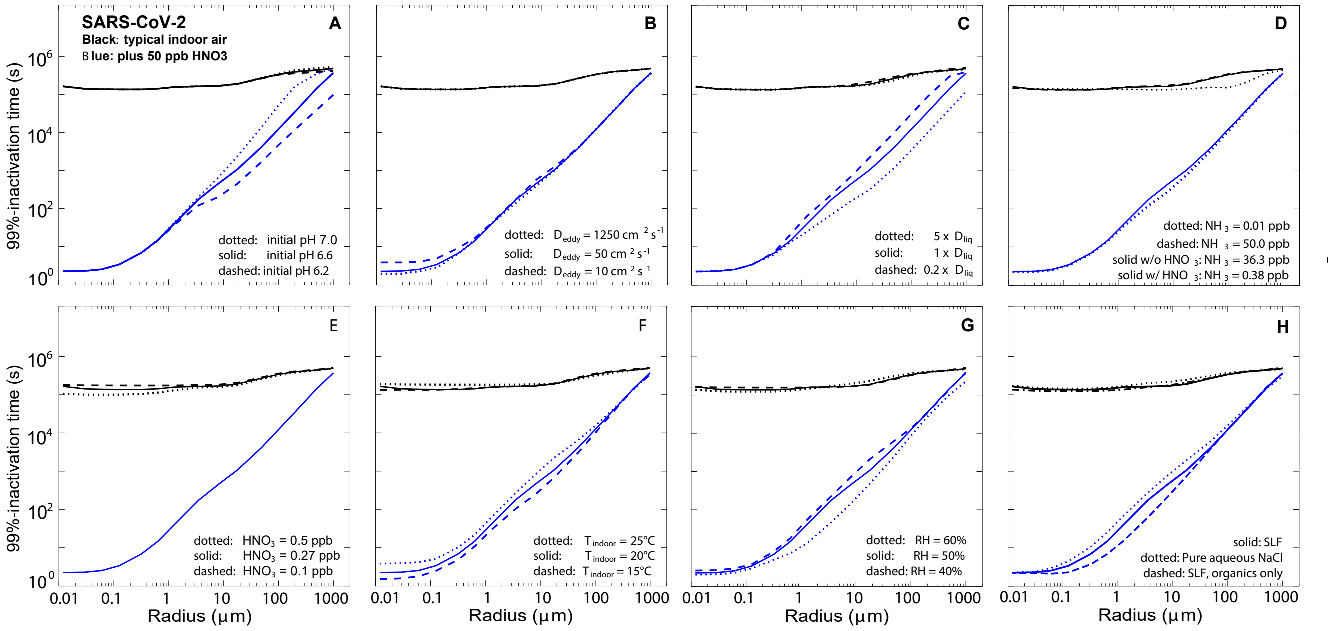


**Fig. S12. Sensitivity studies for the inactivation of SARS-CoV-2.** Same sensitivity tests as in Fig. S11, but for SARS-CoV-2 instead IAV. For Panel **F**, the virus inactivation rates are assumed to increase by a factor of 1.3 when temperature is increased 5°C (following (11)).

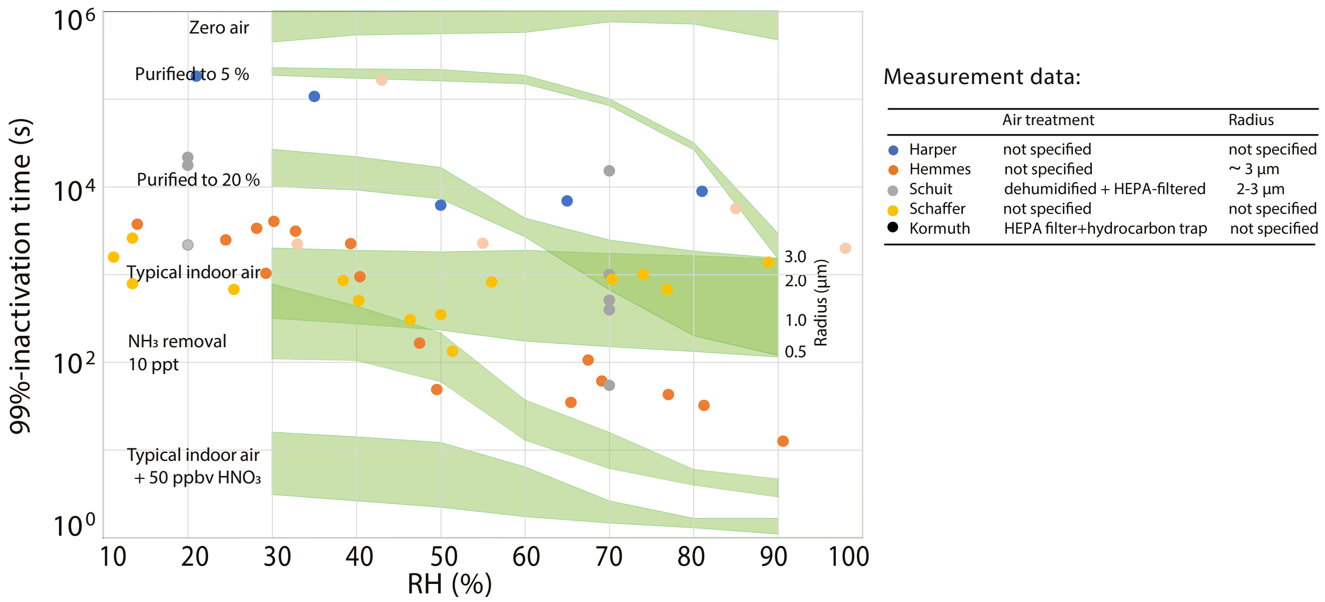


**Fig. S13. Comparison of measured 99%-inactivation times of IAV in aerosols with modeled inactivation for different compositions of air (shaded in green).** Colored circles refer to measurements based on the literature sources listed in the accompanying table (all obtained experimentally at temperatures between 20-25°C). Aerosol particle sizes reported in the literature are typically a few micrometers, but often not specified. Air treatment in the published aerosol experiments, where stated, includes HEPA-filtering and other air purification measures. Such treatments partly remove pH-relevant species from the air, such as $HNO_3$, which increases the aerosol pH (partly compensated by a reduced concentration of $NH_3$). Inactivation times were estimated assuming first-order kinetics, and were calculated from inactivation rate constants, half lives or decay curve data shown in the publications. Areas highlighted in green refer to ResAM model results with aerosol particle radii ranging from 0.5 µm to 3 µm (as indicated on the right side). The inactivation rate itself is a function of time due to the dependence on the changing pH. This implies that cases with 99%-inactivation times of many hours or days are only inaccurately determined by the measurements, which typically last for only about an hour; therefore we constrained the model results to the duration of the experiment and extrapolated the results to achieve measurement-model comparability. The composition of indoor air with and without various treatments used in ResAM is given in Table S4. Citations refer to: Harper (6); Hemmes (5); Schuit (9); Schaffer (7); Kormuth (8). Overlapping data points were slightly shifted in RH for clarity.

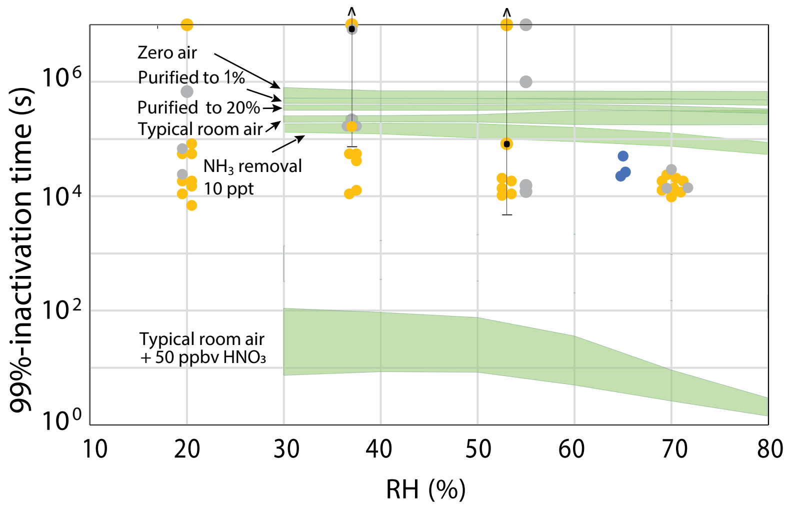


Measurement data:

| | Air treatment | Radius |
|---|---|---|
| ● van Doremalen | not specified | < 2.5 µm |
| ● Dabisch | compressed air | ~ 1 µm |
| ● Schuit | dehumidified + compressed | ~ 1 µm |

**Fig. S14. Comparison of measured 99%-inactivation time of SARS-CoV-2 with modelled inactivation for different composition of air (shaded in green).** Colored circles refer to measurements based on the literature sources listed in the accompanying table (all obtained experimentally at temperatures between 20-23°C). Aside from controlling temperature and RH, air treatment in the published aerosol experiments was not specified, and the air composition under which inactivation data were obtained is therefore unknown. Inactivation times were estimated assuming first-order kinetics, and were calculated from inactivation rate constants or decay curve data shown in the publications. Note that Schuit *et al.* and Dabisch *et al.* also measured inactivation under the influence of solar irradiance, resulting in shorter inactivation times ($\sim 10^3$ s) than the dark inactivation measurements shown here. Measurement variability among experimental replicates is large, in particular at inactivation times larger than one day ($\sim 10^5$ s), as indicated by the arrow shown in two examples. Areas highlighted in green refer to ResAM model results with aerosol particle radii ranging from 0.5 µm to 3 µm (as indicated on the right side). The inactivation rate itself is a function of time due to the dependence on the changing pH. This implies that cases with 99%-inactivation times of many hours or days are only inaccurately determined by the measurements, which typically last for only one to three hours; therefore we constrained the model results to the duration of the experiment and extrapolated the results to achieve measurement-model comparability. The composition of indoor air with and without various treatments used in ResAM is given in Table S4. Citations refer to: van Doremalen (12); Dabisch (11); Schuit (10). Overlapping data points were slightly shifted in RH for clarity.

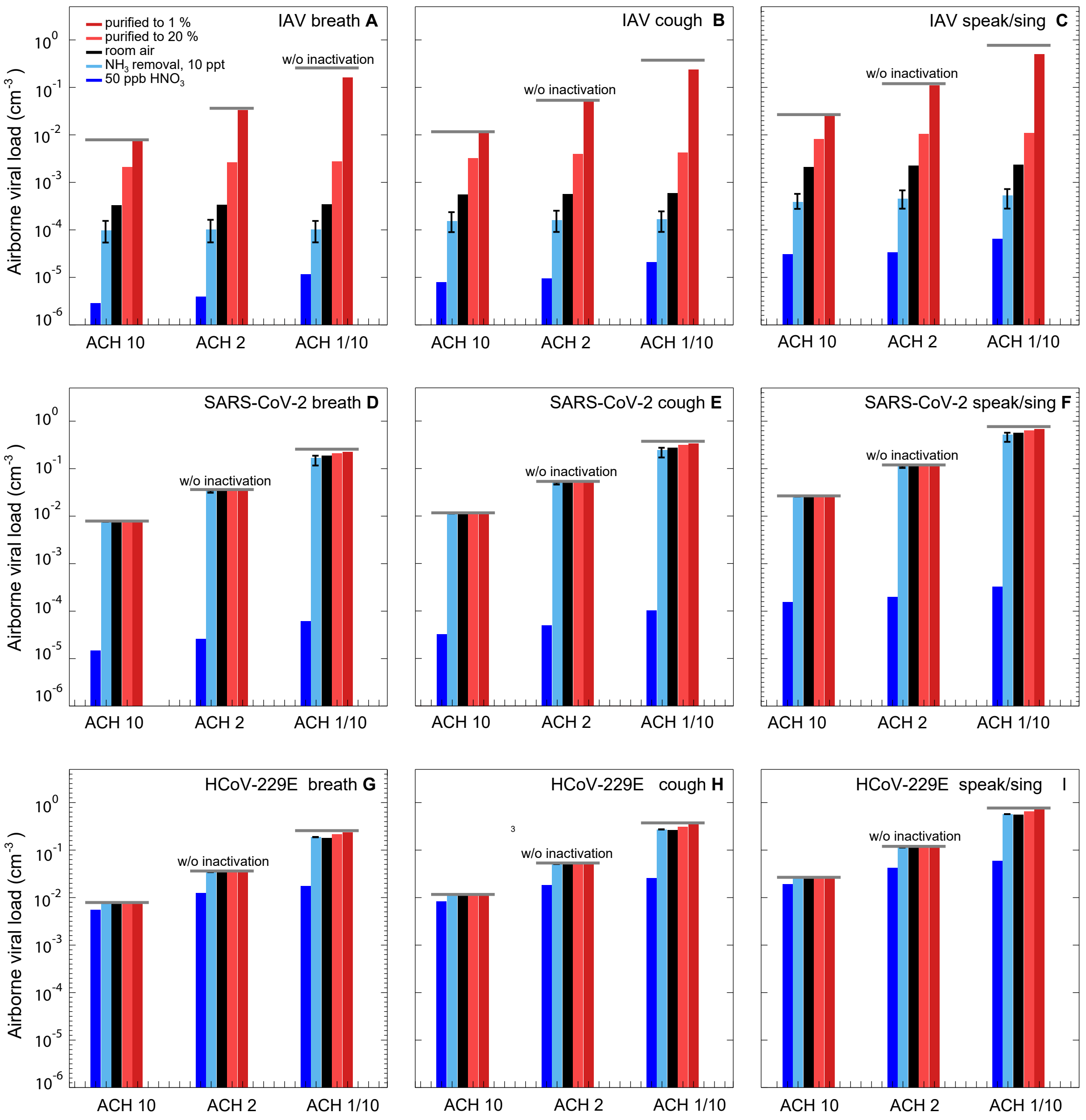


**Fig. S15. Virus loading under steady state conditions in a room with infected persons subject to different ventilation strengths (Air Change per Hour, ACH), and different air treatments for various human activities.** These activities include normal breathing, coughing, and speaking/singing, leading to the exhalation of different sizes of aerosol particles (see Fig. 4E and Ref. (36)). The steady state conditions are calculated assuming one infected person per 10 $m^3$ of air volume, emitting virus-laden aerosol, which is balanced by aerosol removal via ventilation and deposition. Upper row (**A-C**) shows results for IAV, center row (**D-F**) for SARS-CoV-2, and bottom row (**G-I**) for HCoV-229E. Left column (**A**, **D**, **G**) shows results for normal breathing; center column (**B**, **D**, **H**) for coughing; and right column (**C**, **F**, **I**) for speaking or singing. For detailed description see Fig. 5 and supplementary text.

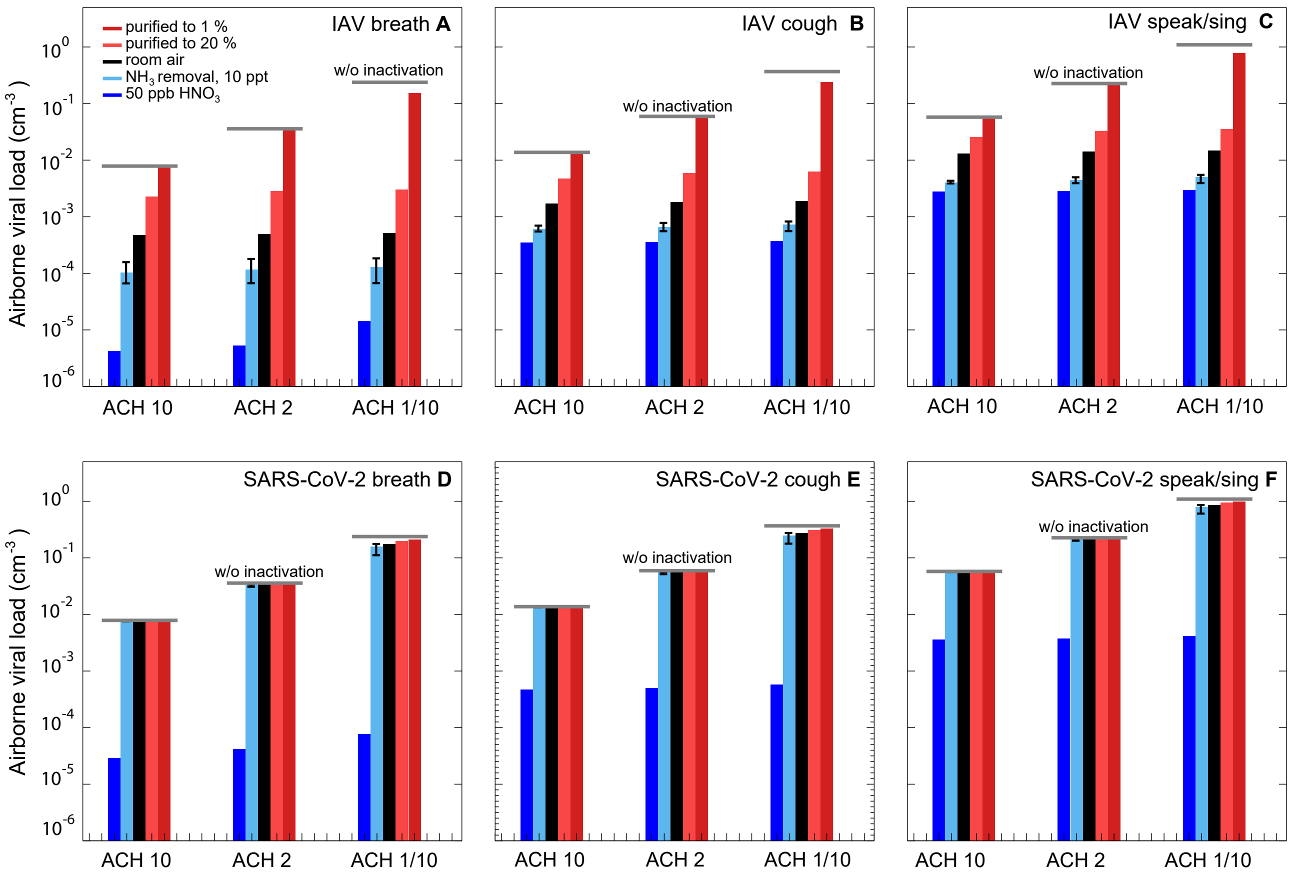


**Fig. S16. Steady state virus loading at different ACH and different room conditions for breathing, coughing, speaking or singing.** Same as Fig. S15 for IAV and SARS-CoV-2, but assuming that the virus concentration is proportional to the size of the aerosol particles (as motivated by data assembled by Poehlker et al. (36)).

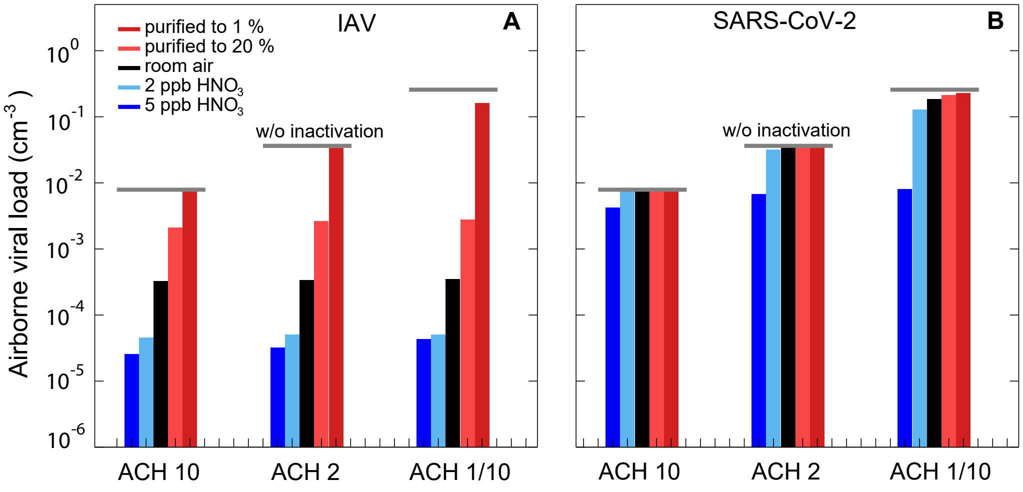


**Fig. S17. Steady state IAV and SARS-CoV-2 airborne viral load, specifically for air treatment with very low $HNO_3$ concentrations.** Same as Fig. S15A+D, but using only 2 and 5 ppb $HNO_3$ to acidify the indoor air.

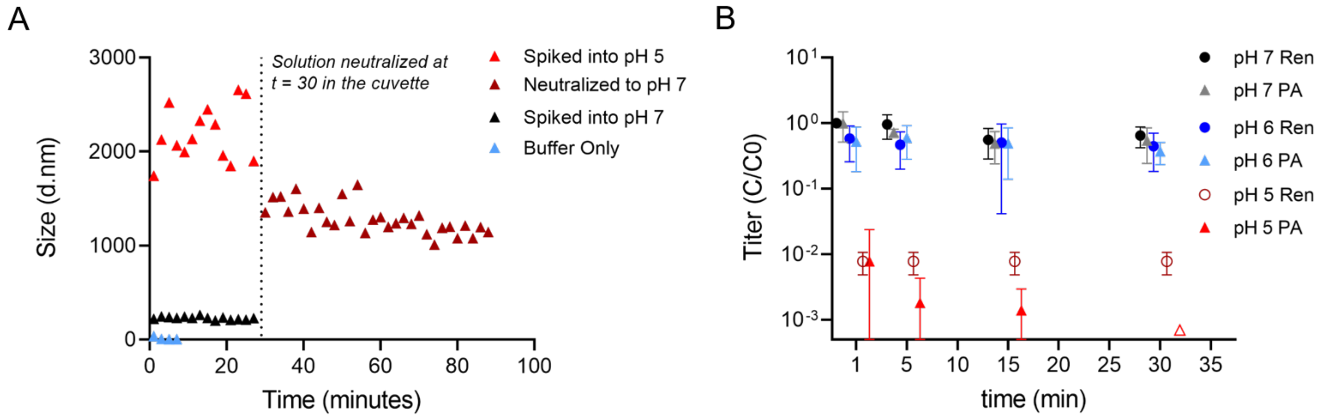


**Fig. S18. Effect of virus aggregation on the infectivity of IAV.** (**A**) IAV particle diameters were measured by DLS in PBS adjusted to pH 7 or pH 5. After 30 min the latter was neutralized to pH 7. The average diameter was measured every 2 min over a total time of 90 min. At pH 7, the average particle size was stable and corresponds to the diameter of monodisperse IAV particles (between 100 - 200 nm). The average particle diameter immediately increased to 1800 nm at pH 5, indicating rapid aggregation. The mean diameter then fluctuated between 1900 – 2600 nm for the full 30 minute period, indicating these aggregates were not transient. Upon neutralization to pH 7, aggregation was only partly reversed and the mean particle diameter remained above 1000 nm for the remainder of the time-course. (**B**) Infectivity of WSN/33-Renilla IAV particles over time in PBS adjusted to either pH 7, pH 6 or pH 5. WSN/33-Renilla harboring a gene encoding Renilla luciferase instead of the HA gene was generated as previously described (44). The titer was determined by both plaque assay and Renilla luciferase assay. While the titer measured by plaque assay (PA) indicates the number of infectious units, the titer measured by Renilla assay (Ren) correlates with gene expression. Both plaque and Renilla assay indicate the same decrease in infectivity as a function of pH, confirming that virus titer loss is associated with inactivation (i.e., loss in viral gene expression), not solely aggregation. Values below the LoD are indicated with empty symbols. Residual titer fractions were derived by normalization of measured titers ($C$) to the initial titer ($C_0$).

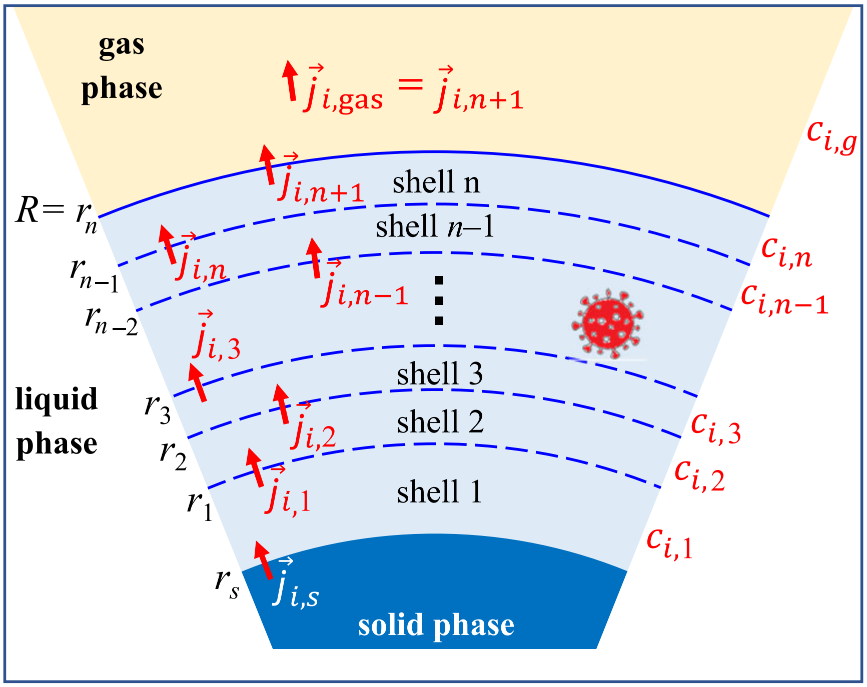


**Fig. S19. Illustration of the discretization scheme used in the biophysical diffusion model ResAM.** A spherical particle of radius $R$ is divided into $n$ concentric liquid shells plus a solid salt core of radius $r_s$ (if efflorescence occurred). In each time step, changes in the concentration $C_{i,k}(t)$ of species $i$ in shell $k$ are calculated by fulfilling the liquid phase diffusion equation (Eq. S7), i.e. from the continuity equation balancing the diffusion fluxes $\vec{j}_{i,k}(t)$ into and out of the shell with concentration changes in the shell. The flux in the gas phase fulfills the gas phase diffusion equation. The flux into or out of the particle is calculated by equating $\vec{j}_{i,n+1}(t)$ with the flux in the gas phase of species $i$ above the particle surface. The flux from or to the solid core, $\vec{j}_{i,s}(t)$, is calculated using Eq. S17.

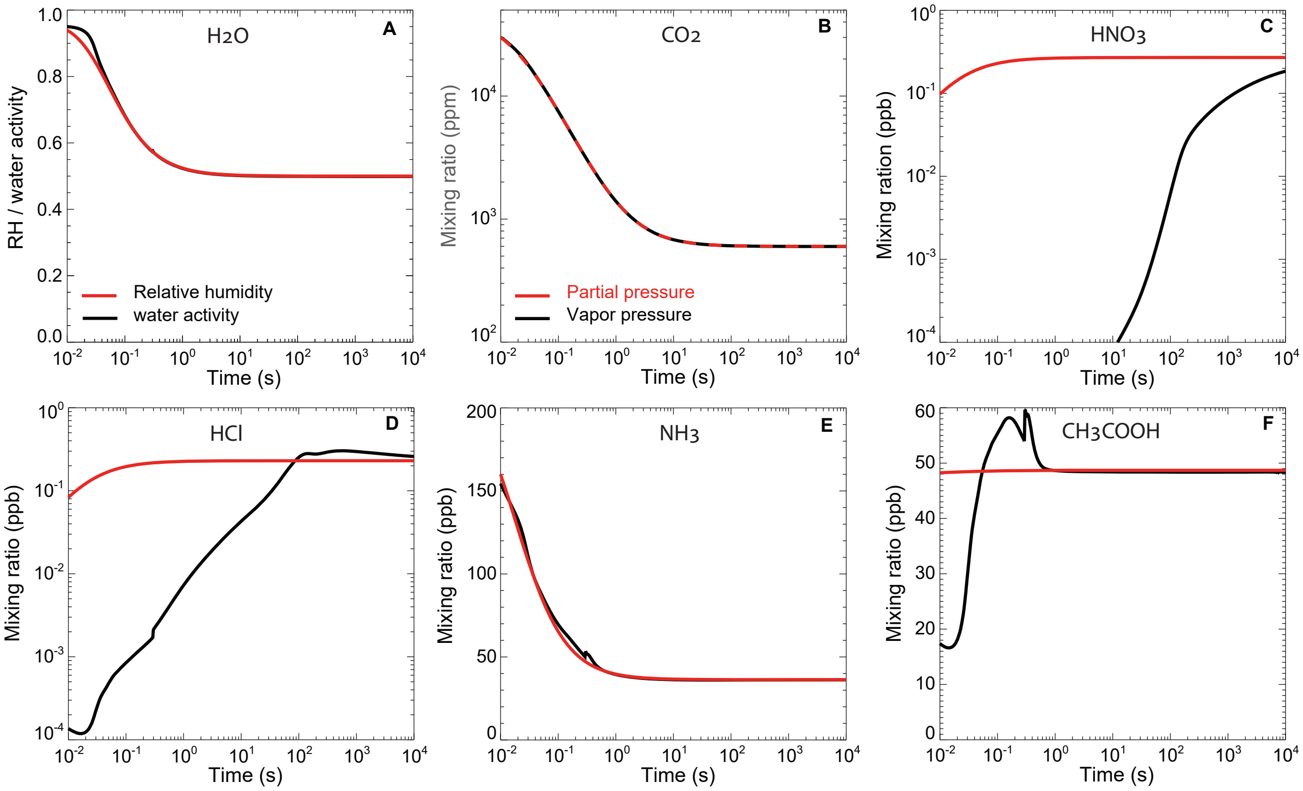


**Fig. S20. Non-equilibrium conditions of chemical species in the gas phase above expiratory particle surface leading to their growth or evaporation.** Relative humidity and mixing ratios (or mole fractions) of gaseous species in the exhaled plume mixing with ambient air (red lines) in comparison with equilibrium values corresponding to the vapor pressures of the exhaled particles (black lines) for typical indoor air simulations shown in Fig. 3. Panel (**A**) shows relative humidity and water activity of the outermost shell; panels (**B**) to (**F**) show $CO_2$, $HNO_3$, HCl, $NH_3$, and $CH_3COOH$. The difference between the red and black lines represents the force on the particles, i.e. supersaturation and uptake of the gaseous species when the red values are higher, subsaturation and loss of the species to the gas phase when the black values are higher.

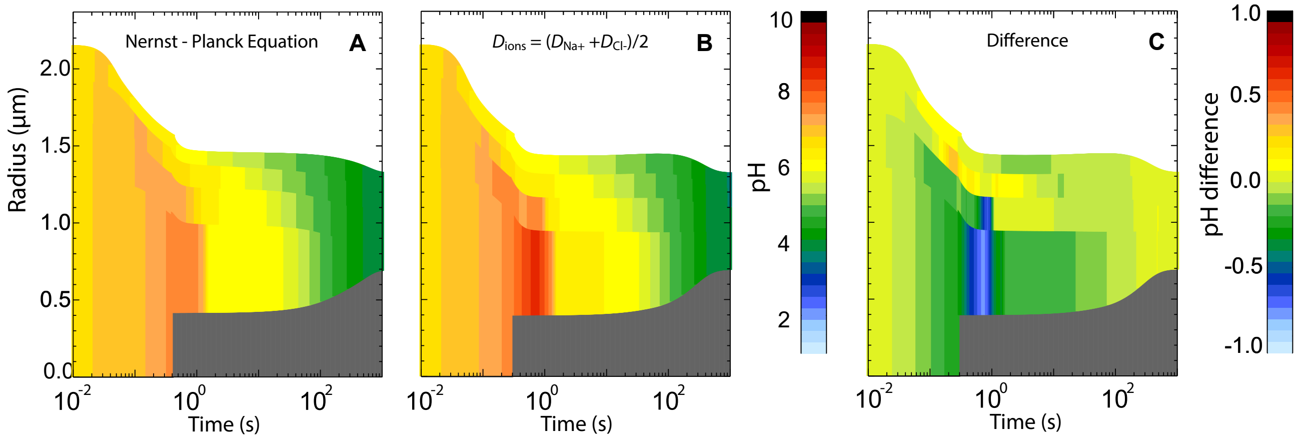


**Fig. S21. Comparison of a treatment of diffusion and electroneutrality using the Nernst-Planck equation with a standard treatment using Fick's Law.** (**A**) Nernst-Planck treatment used in the present work based on activity gradients in Eq. S6. (**B**) Treatment assuming all ions to have the same diffusion coefficient in SLF and solving the diffusion equation with Fick's law based on concentration gradients. (**C**) Difference of (A) minus (B), showing that large pH differences can occur in the vicinity of large activity of concentration gradients.

### Supplementary Tables

#### Table S1. Composition of the synthetic lung fluid (SLF), adapted from Bicer (45)

| **Components** | **Concentration (mass per volume of solution)** |
|---|---|
| **Solvent** | |
| Water | - |
| **Salts** | |
| Calcium chloride dihydrate | 0.185 mg/ml |
| Magnesium sulfate heptahydrate | 0.200 mg/ml |
| Potassium chloride | 0.400 mg/ml |
| Potassium phosphate monobasic anhydrous | 0.060 mg/ml |
| Sodium bicarbonate | 0.350 mg/ml |
| Sodium chloride | 8.000 mg/ml |
| Sodium phosphate dibasic heptahydrate | 0.090 mg/ml |
| **Sugars** | |
| Dextrose anhydrous | 1.000 mg/ml |
| **Proteins** | |
| Albumin | 8.800 mg/ml |
| Transferrin | 1.500 mg/ml |
| **Lipids** | |
| DPPC | 4.800 mg/ml |
| DPPG | 0.500 mg/ml |
| Cholesterol | 0.100 mg/ml |
| **Antioxidants** | |
| Ascorbic acid | 0.025 mg/ml |
| Uric acid | 0.016 mg/ml |
| GSH (glutathione) | 0.052 mg/ml |

**Table S2. Equilibrium constants used for the kinetic simulations. They are valid for environment temperature from 0°$C$ to 40°$C$.**

| Reaction | | | Equilibrium constant | Unit | Source |
|---|---|---|---|---|---|
| $H_2O_{(aq)}$ | $\rightleftharpoons$ | $H_2O_{(gas)}$ | $p_{H_2O} = a_w \times p_{\text{pure water}}$, [a] | Pa | (58, 59, 77) |
| $H_2O_{(aq)}$ | $\rightleftharpoons$ | $H^+ + OH^-$ | $\exp\left(-9.2644 - \frac{6872.7}{T}\right)$ | molality$^2$ | (89) |
| $H^+ + NH_{3(gas)}$ | $\rightleftharpoons$ | $NH_4^+$ | $\exp\left(-5.2167 + \frac{9826.67}{T} - 0.00787 \times T\right)$ | Atm$^{-1}$ | (90) |
| $CH_3COOH_{(gas)}$ | $\rightleftharpoons$ | $CH_3COOH_{(aq)}$ | $4000 \times \exp\left(6200\left(\frac{1}{T} - \frac{1}{298.15}\right)\right)$ | molality Atm$^{-1}$ | (91) |
| $CH_3COOH_{(aq)}$ | $\rightleftharpoons$ | $H^+ + CH_3COO^-$ | [b] | molality | (92) |
| $HCl_{(gas)}$ | $\rightleftharpoons$ | $H^+ + Cl^-$ | [c] | molality$^2$Atm$^{-1}$ | (58, 59) |
| $HNO_{3(gas)}$ | $\rightleftharpoons$ | $H^+ + NO_3^-$ | [d] | molality$^2$Atm$^{-1}$ | (58, 59) |
| $CO_{2(aq)} + H_2O_{(aq)}$ | $\rightleftharpoons$ | $H^+_{(aq)} + HCO_3^-$ (aq) | $4.448 \times 10^{-7} \exp(-2133(\frac{1}{T} - \frac{1}{298.15}))$ | molality | (93, 94) |
| $HCO_3^-$ (aq) | $\rightleftharpoons$ | $H^+_{(aq)} + CO_3^{-2}$(aq) | $1.08555 \times 10^{-9} \exp(-3347.3(\frac{1}{T} - \frac{1}{298.15}))$ | molality | (94) |
| $CO_{2(gas)}$ | $\rightleftharpoons$ | $CO_{2(aq)}$ | $0.034 \times \exp\left(2300\left(\frac{1}{T} - \frac{1}{298.15}\right)\right)$ | molality Atm$^{-1}$ | (91) |
| $NH_4CH_3COO_{(ad)}$ | $\rightleftharpoons$ | $NH_4^+{}_{(aq)} + CH_3COO^-{}_{(aq)}$ | $3.44 \times 10^{-3}$ at $T = 293.55$ K only | molality | [e] |
| $NaCl_{(solid)}$ | $\rightleftharpoons$ | $Na^+ + Cl^-$ | $13.812 - 0.025681 \times (T - 298.15)$ | molality$^2$ | [f] |

[a] $p_{\text{pure water}}$ is given by Equation (11) of Murphy and Koop (77)

[b] $\log K = -\frac{1500.65}{T} - 6.50923 \log T - 0.0076792 \times T$

[c] $\ln K = 14.53335 + 3067.38(\frac{1}{T} - \frac{1}{298.15}) - 19.91 \ln(\frac{T}{298.15}))$

[d] $\ln K = 394.007 - \frac{3020.3522}{T} - 71.002 \ln(T) + 0.131442311 \times T - 0.420928363 \times 10^{-4} \times T^2$.

[e] The dissociation coefficient of $NH_4CH_3COO$ is estimated from our own measurement of $NH_3$ vapor which is 0.12 Pa at $T = 293.55$ K and RH of 87.5%.

[f] The activity product of solid NaCl is calculated with ResAM using the NaCl solubility data in water (95).

**Table S3. Liquid phase diffusion coefficients of chemical species relevant in expiratory aerosol particles in infinitely diluted water at 298.15 K.**

| Species | $D_\ell$ in $10^{-5}$ cm$^2$s$^{-1}$ | Source | Species | $D_\ell$ in $10^{-5}$ cm$^2$s$^{-1}$ | Source |
|---|---|---|---|---|---|
| $H_2O$ | 2.44 | (96) | $NH_3$ | 1.7 | (97) |
| $H^+$ | 9.3 | (96) | $NH_4^+$ | 1.7 | (98) |
| $Na^+$ | 1.33 | (96) | $CH_3COOH$ | 1.18 | (67) |
| $Cl^-$ | 2.03 | (96) | $CH_3COO^-$ | 1.08 | (67) |
| $NO_3^-$ | 1.90 | (96) | $NH_4CH_3COO$ | 1.18 | *a* |
| $OH^-$ | 5.25 | (96) | $CO_2$ | 1.98 | (97) |
| Proteins/lipids | 0 | *b* | $HCO_3^-$ | 1.11 | (97) |

[a] We assume the molecular form of ammonium acetate, $NH_4CH_3COO$, to have the same diffusion coefficient as $CH_3COOH$.

[b] The liquid phase diffusion of lipids and proteins is assumed to be extremely slow and can be ignored.

**Table S4. Trace gases in exhaled air and in indoor air for different scenarios. In the present study, we assume the relative humidity of the indoor air to be 50 % and the temperature 293.15 K, unless stated differently (e.g., Fig. S8 and Figs. S11E,G and S12E,G). The background aerosol loading is assumed to be 20 μg/m$^3$. The temperature of exhaled air at time zero is assumed to be 307.15 K (75). The entries with enriched acetic acid ($CH_3COOH$) serve as basis for Fig. S9.**

| Scenario | RH | $NH_3$/ppb | $HNO_3$/ppb | HCl/ppb | $CH_3COOH$/ppb | $CO_2$/ppm |
|---|---|---|---|---|---|---|
| exhaled air | 91%[a] | $238 \pm^{500}_{200}$ [b] | 0.0[c] | 0.0[c] | 486(99, 100) | 46'500[d] |
| Typical indoor air | 50% | $36.3 \pm 35$ [e] | $0.27 \pm 0.03$ [f] | 0.23(25) | 48.7(25) | 600(25) |
| Purified indoor air to 20% | 50% | 7.26 | 0.054 | 0.046 | 9.74 | 600 |
| Purified indoor air to 1% | 50% | 0.363 | 0.003 | 0.0023 | 0.487 | 600 |
| Zero air | 50% | 0 | 0 | 0 | 0 | 600 |
| Indoor air, $NH_3$ removal | 50% | 0.01[g] | $0.53\pm^{0.67}_{0.26}$ [h] | 0.23 | 48.7 | 600 |
| Indoor air + 50 ppb $HNO_3$ | 50% | 0.38[g] | 50 | 0.23 | 48.7 | 600 |
| Indoor air + 5 ppm $CH_3COOH$ | 50% | 35.8[g] | 0.27 | 0.23 | $5\times10^3$ | 600 |
| Indoor air + 50 ppm $CH_3COOH$ | 50% | 184.7[g] | 0.93 | 0.23 | $50\times10^3$ | 600 |
| Indoor air + 100 ppm $CH_3COOH$ | 50% | 11.10[g] | 0.43 | 0.23 | $100\times10^3$ | 600 |

[a] Berry reported a $H_2O$ amount in exhaled air of 0.0342 grams per liter (75). A relative humidity of 91% is obtained using the vapor pressure expression of Murphy and Koop (77) at exhalation temperature.

[b] See Nazaroff and Weschler (25), page 575.

[c] There are no literature values for HCl and $HNO_3$ concentrations in exhaled air. Thus, we assume they are zero.

[d] $CO_2$ concentration in exhaled air is $(46.5\pm6.5)\times10^3$ ppm (101).

[e] Weighted average over all values in Table 6 of Nazaroff and Weschler (25).

[f] Weighted average over all values with natural ventilation or window air conditioners in Table 12 of Nazaroff and Weschler (25).

[g] When adding $HNO_3$ to indoor air, ammonia is reduced due to the uptake by the background aerosol. Prior to the addition, the indoor air is assumed to have a loading of 20 μg/m$^3$. The molalities of indoor background aerosol (with water activity of 0.5 corresponding to 50% RH) are: 6.36 mol/kg sucrose, 6.36 mol/kg sulfate ions, 23.89 mol/kg $NH_4{}^+$, and 11.16 mol/kg $NO_3{}^-$ .

[h] When $NH_3$ is scrubbed from the indoor air while the particulate matter is not removed, the ammonium ($NH_4^+$) in the background particles will partition to the gas phase. This results in the release of $HNO_3$ to the gas phase from the nitrate ($NO_3^-$) in the particular matter, enhancing the virus inactivation. The lower $HNO_3$ limit specified here refers to zero $NO_3^-$ in the background aerosol and the upper limit to 40 nmol/m$^3$ $NO_3{}^-$, as summarized by (25).
